# Germline sequencing in children with cancer in Quebec: an integrated investigative approach

**DOI:** 10.64898/2026.02.10.26345751

**Authors:** Edith Sepulchre, Alexandre Rouette, Claire Freycon, Leora Witkowski, Safa Jammali, Thomas Sontag, Sylvie Langlois, Noémie Sutan, Crystal Budd, Véronique Lisi, Chantal Richer, Loubna Jouan, Marie-Ève Lepage, Lara Reichman, William Foulkes, Anne-Marie Laberge, Bruno Michon, Josée Brossard, Nada Jabado, Daniel Sinnett, Thai-Hoa Tran, Stéphanie Vairy, Raoul Santiago, Sonia Cellot, Catherine Goudie, Vincent-Philippe Lavallée

**Author notes:** Co-corresponding authors: Contact information, Catherine Goudie, MD MSc, Vincent-Philippe Lavallée, MD PhD. co-senior authors (Catherine Goudie, Vincent-Philippe Lavallée).

## Abstract

**Background:** The province of Quebec has progressively implemented paired tumour-germline sequencing in paediatric oncology through two coordinated precision research programs, preceding a province-wide mainstream clinical genomics initiative. We report the prevalence, spectrum, and clinical relevance of germline findings (GFs) in children with primary extracranial cancers, integrating molecular, phenotypic, and pathological data.

**Methods:** Patients enrolled between 2014 and 2022 underwent germline whole-exome sequencing (WES) using a virtual 352-cancer gene panel. Sequencing, bioinformatics and variant interpretation followed best practices standards based on GATK, ACMG/AMP and ClinGen recommendations. Somatic WES and transcriptomic data were integrated when available. GFs were categorised as diagnostic findings (DFs; established or suspicious association with the cancer phenotype) or as other findings further subcategorised according to actionability and age of disease onset.

**Findings:** Among 484 children, 130 (26.9%) carried 149 GFs, including 49 (10.1%) with a DF (42 with well-established associations with cancer phenotypes). DFs involved 21 genes related to childhood cancer predisposition, trisomy 21 and one clinical Beckwith-Wiedemann syndrome. Six DFs were initially missed by standard exome pipelines, and mosaic constitutional cancer predisposition syndrome (CPS) was confirmed in 4/49 children, underscoring the value of integrative analyses. A CPS was known at the time of primary cancer in 10/49 children. Among those diagnosed with a CPS after cancer onset, suggestive phenotypic features were present in 36/39. Other non-diagnostic findings were identified in 92 children; 21 (4.3% of the cohort) with actionable implications in childhood (n=7) or adulthood (n=14). Somatic sequencing was informative for refining causality, as somatic second hit alterations were identified in 29/33 (87.9%) DFs involving monoallelic tumour suppressor genes, whereas no such alterations were observed in non-DFs counterparts (0/57; p<0.0001). **Interpretation:** This provincial research experience highlights the analytical and practical challenges of germline evaluation in paediatric oncology and supports a shift toward integrative interpretation frameworks combining complementary germline, somatic, pathology, and phenotypic data. Flexibility in investigative strategies and nuanced categorisation of findings are warranted, guided by a child-centred interpretative framework. This approach underpins Quebec’s paediatric oncology genomics mainstreaming initiative.

## INTRODUCTION

Pathogenic/likely pathogenic germline variants (PGVs) in over 150 different cancer predisposing genes (CPGs) are known to contribute to over 50 paediatric cancer predisposition syndromes (CPS) (1). Identifying a CPS enables tailored management, targeted surveillance, risk-reduction strategies, and identification of at-risk family members.

Until 2015, a primarily phenotype-driven approach, relying on clinical suspicion and collaboration with genetics services, was the main diagnostic pathway for identifying CPS. Since the landmark publication by Zhang et al in 2015 (2), more than 25 paediatric cancer sequencing initiatives have reported CPS frequencies ranging from 3.8 to 18% among children diagnosed with cancer (3,4). Published studies differ in cohort inclusion criteria, breadth and content of gene panels, sequencing pipelines, as well as in definitions and thresholds applied to classify germline findings (GFs) as “positive” or “negative”. Such heterogeneity explains the wide range of reported diagnostic yields for CPS and highlights uncertainties regarding (i) the optimal scope of gene panels (balancing the risk of missing relevant variants against the inclusion of genes with limited clinical utility), (ii) the strength and specificity of gene-cancer associations, and (iii) the interpretation of inheritance patterns, particularly for heterozygous variants in recessive or adult-onset CPS genes. Despite these challenges, paediatric oncology sequencing initiatives provide a strong rationale for some institutions and jurisdictions to adopt comprehensive germline DNA sequencing as part of routine clinical care (genotype-first approach).

The province of Quebec (Canada) has been at the forefront of integrating genomics into paediatric oncology care, evolving from research-based paired tumour-germline sequencing in patients with hard-to-treat malignancies (since 2014), to universal paired DNA sequencing and transcriptome analysis for all children with cancer (since 2018). This comprehensive research program paved the way for the implementation, in 2023, of publicly funded clinical paired tumour-germline sequencing and transcriptome analysis for all children with cancer in Quebec, with research approaches strategically complementing the funded clinical investigations(5).

This study outlines the practical lessons learned from precision oncology research in Quebec, focusing on GFs correlated with somatic analyses in this population. Through phenotype-genotype correlation analyses in 484 children with primary non-central nervous system (CNS) cancers, we delineate both the strengths and limitations of current CPS diagnostic strategies in real-world clinical and research settings.

## MATERIAL AND METHODS

### Study Design and Patients

Paediatric oncology patients from Québec were enrolled onto one or both precision oncology research programs, based on year of enrollment and/or cancer type (figure S1). The “Triceps” program (2014–2022) included patients with hard-to-treat malignancies, mostly relapsed/refractory patients, while the “Signature” initiative, launched in 2019 (and ongoing), includes all newly diagnosed or relapsed paediatric oncology patients irrespective of cancer type or prognosis. Importantly, both programs involved identical paired tumour-germline whole-exome sequencing (WES) and somatic transcriptome workflows. While patients with primary CNS tumours were eligible for both research initiatives, most underwent alternative research sequencing approaches. For this reason, we excluded patients with primary CNS tumours in this study population. Written informed consent was obtained for all participants. Both studies were approved by the institutional Research Ethics Boards of the four paediatric oncology hospitals in Quebec, with CHU Sainte-Justine as the lead institution. All participants underwent germline WES. Bioinformatic processing was performed using Genome Analysis Toolkit (GATK) pipeline.

### Data Collection

Clinical information was collected through dedicated questionnaires and electronic medical chart review (figure S2). The following data were systematically extracted: sex, age, anthropometric measurements at cancer diagnosis, International Classification of Diseases (ICD) codes, tumour characteristics, pathology findings (immunohistochemistry [IHC], karyotyping, micro-array, fluorescence in situ hybridization), and disease course. Clinical features, including growth, neurodevelopment, congenital anomalies, relevant laboratory results, family history of cancer were collected, along with clinical genetic consultations and testing results.

### Variant Prioritization and Classification

Details regarding sampling, sequencing, data processing, and variant curation are provided in Supplementary Methods. Annotated variants from germline WES were filtered to retain only those located in a predefined virtual panel of 352 cancer-related genes (table S1). Variants classified as pathogenic/likely pathogenic in Clinvar with a review status of “reviewed by expert panel” (three stars) were accepted as such. Remaining variants were filtered based on ClinVar annotation, variant type, prediction scores, and minor allele frequency in gnomAD. Variants were then curated following ACMG 2015 guidelines (6), complemented by ClinGen gene-specific variant curation expert panels recommendations when available(7), and by selected adaptations from recent expert recommendations (table S2A–B, figure S2). Variants annotated as pathogenic/likely pathogenic were retained and are herein collectively referred to as PGVs. Variants conferring reduced penetrance and risk alleles were curated in line with recent consensus frameworks (8,9). Variants of uncertain significance (VUS) were retained when supported by strong molecular evidence for tumourigenesis and clinical evidence for association with cancer phenotype, referred to hereafter as VUS+. In this study, GFs include all PGVs, reduced penetrance/risk alleles, VUS+ (filtered gene panel), as well as chromosomal anomalies (karyotype/microarray). To fully characterise our paediatric oncology cohort regardless of genetic diagnosis method, we also classified any clinically confirmed CPS diagnosis by a medical geneticist as a GF. Somatic WES and transcriptomic data were also considered, when available, to refine germline variant classification or confirm mosaic and splicing effects (Supplementary Methods).

### Categorisation of GFs by disease relevance and actionability

GFs were then classified into (1) Diagnostic findings (DF), defined by an established or highly suspicious association with the phenotype, and (2) Other findings, encompassing GFs not known to be associated with the phenotype (figure 1). Other findings were then subclassified into actionable other findings in paediatric age and/or adulthood, non-actionable other findings, carrier status for autosomal recessive disorders, and low-penetrance/risk alleles. DFs and actionable other findings in paediatric age were disclosed to participants, while carrier statuses and low-penetrance/risk alleles were not disclosed. Actionable findings restricted to adulthood, as well as findings with likely no actionability, were disclosed on a case-by-case basis, in accordance with participant consent and clinical and study context. Genetic results were disclosed as per research and institutional clinical procedures, and patients were referred to clinical genetics for counseling and subsequent management.

**Figure 1.**
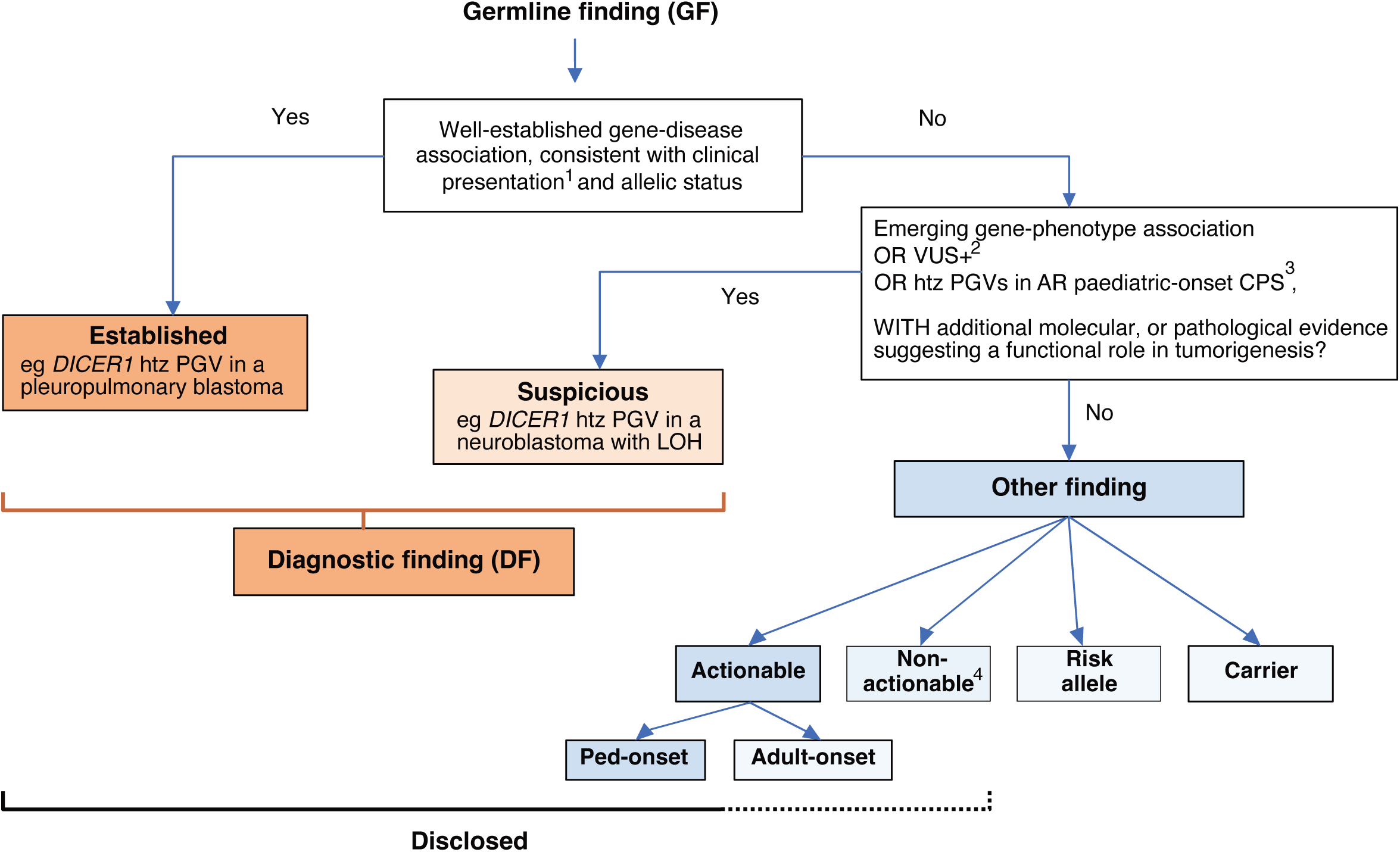
Decision tree for the categorisation of germline findings by disease relevance. Decision tree illustrating the classification of germline findings (GFs) into diagnostic findings (DF) and other findings, based on the strength of the gene-disease association and consistency with the clinical presentation. Diagnostic findings include both established and suspicious associations supported by suggestive but incomplete evidence. Other findings were further subclassified into: (a) actionable other findings in paediatric age and/or adulthood, (b) likely non-actionable other findings, (c) carrier status for autosomal recessive disorders, and (d) low-penetrance or risk alleles. Notes: 1: Clinical presentation includes tumour type, age of onset, clinical features, and personal and family history. 2: Gene associated with the child’s phenotype. 3: Genes typically requiring biallelic alteration for paediatric-onset cancer predisposition syndromes. 4: No cancer surveillance recommendation or change in management plan. **Abbreviations:** AR, autosomal recessive; CPS, cancer predisposition syndrome; DF, diagnostic finding; GF, germline finding; htz, heterozygous; LOH, loss of heterozygosity; PGV, pathogenic/likely pathogenic variant; VUS+, highly suspicious variant of uncertain significance.

## RESULTS

### Patient Demographics

Germline WES was performed on 484 paediatric patients with non-CNS primary cancers (table S3). Somatic WES and transcriptome analyses were performed in 470 (97%) and 427 (88%) patients, respectively. The median age at first cancer diagnosis was 7.7 years (range: 0-18.9), and the cohort showed a male predominance (280/484, 58%) (table 1). Most patients (n=351, 72.5%) were enrolled at the time of their first cancer diagnosis. Haematological malignancies accounted for 239/484 (49.5%), with most frequent subtypes including B-cell acute lymphoblastic leukemia (B-ALL) (n=138), acute myeloid leukemia (AML; n=47), and T-cell acute lymphoblastic leukemia (T-ALL) (n=31). Solid tumours made up the remaining 50.5%, with neuroblastoma (n=46), osteosarcoma (n=33), Wilms tumours (n=27), Ewing sarcoma (EWS, n=26) and rhabdomyosarcoma (n=22) being the most frequent diagnoses. A CPS diagnosis was already known in 10 (2%) children at the time of first cancer diagnosis and consisted of trisomy 21 (T21; n=5), ataxia-telangiectasia (A-T; n=1), *APC*-associated polyposis (n=1), BWS (n=1), neurofibromatosis type 1 (n=1) and Rubinstein-Taybi syndrome (n=1).

**Table 1.**
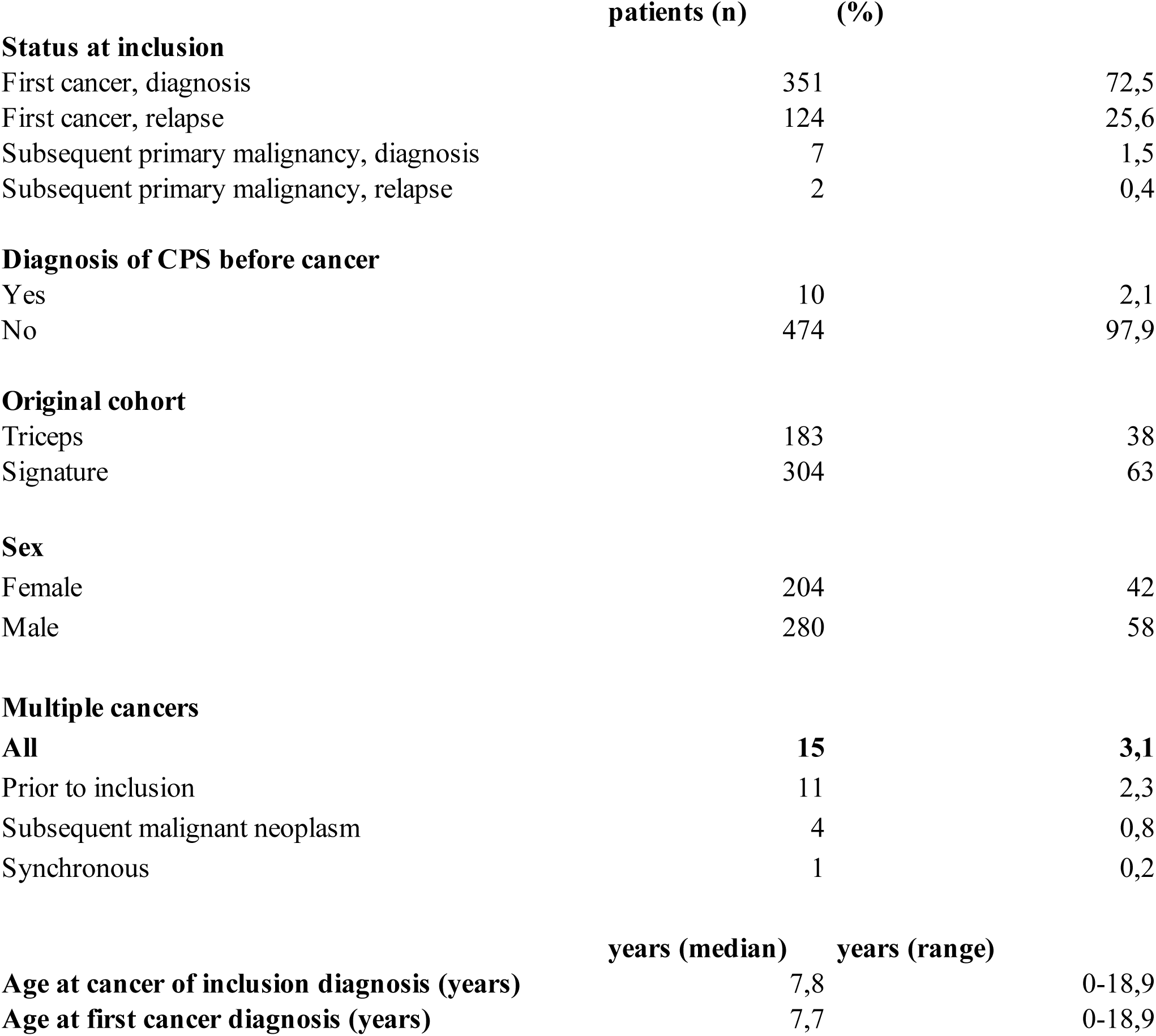
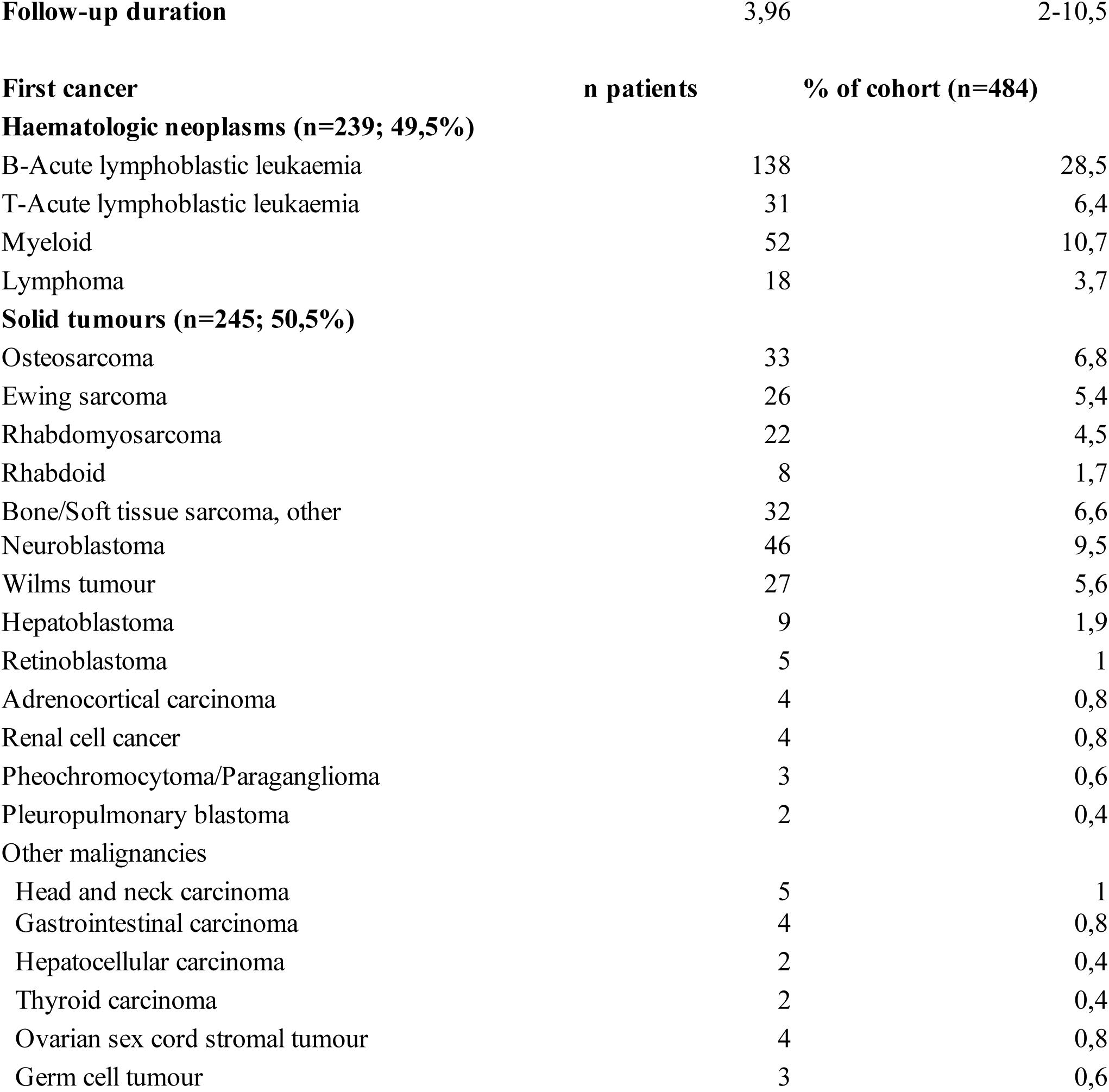
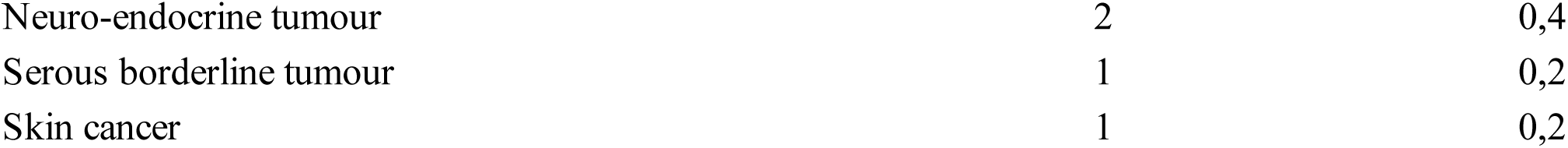
Demographics. Abbreviations: CPS, cancer predisposition syndrome.

|  | patients (n) | (%) |
| --- | --- | --- |
| **Status at inclusion** |  |  |
| First cancer, diagnosis | 351 | 72,5 |
| First cancer, relapse | 124 | 25,6 |
| Subsequent primary malignancy, diagnosis | 7 | 1,5 |
| Subsequent primary malignancy, relapse | 2 | 0,4 |
| **Diagnosis of CPS before cancer** |  |  |
| Yes | 10 | 2,1 |
| No | 474 | 97,9 |
| **Original cohort** |  |  |
| Triceps | 183 | 38 |
| Signature | 304 | 63 |
| **Sex** |  |  |
| Female | 204 | 42 |
| Male | 280 | 58 |
| **Multiple cancers** |  |  |
| **All** | **15** | **3,1** |
| Prior to inclusion | 11 | 2,3 |
| Subsequent malignant neoplasm | 4 | 0,8 |
| Synchronous | 1 | 0,2 |
|  | **years (median)** | **years (range)** |
| **Age at cancer of inclusion diagnosis (years)** | 7,8 | 0-18,9 |
| **Age at first cancer diagnosis (years)** | 7,7 | 0-18,9 |
| **Follow-up duration** | 3,96 | 2-10,5 |
| **First cancer** | **n patients** | **% of cohort (n=484)** |
| **Haematologic neoplasms (n=239; 49,5%)** |  |  |
| B-Acute lymphoblastic leukaemia | 138 | 28,5 |
| T-Acute lymphoblastic leukaemia | 31 | 6,4 |
| Myeloid | 52 | 10,7 |
| Lymphoma | 18 | 3,7 |
| **Solid tumours (n=245; 50,5%)** |  |  |
| Osteosarcoma | 33 | 6,8 |
| Ewing sarcoma | 26 | 5,4 |
| Rhabdomyosarcoma | 22 | 4,5 |
| Rhabdoid | 8 | 1,7 |
| Bone/Soft tissue sarcoma, other | 32 | 6,6 |
| Neuroblastoma | 46 | 9,5 |
| Wilms tumour | 27 | 5,6 |
| Hepatoblastoma | 9 | 1,9 |
| Retinoblastoma | 5 | 1 |
| Adrenocortical carcinoma | 4 | 0,8 |
| Renal cell cancer | 4 | 0,8 |
| Pheochromocytoma/Paraganglioma | 3 | 0,6 |
| Pleuropulmonary blastoma | 2 | 0,4 |
| Other malignancies |  |  |
| Head and neck carcinoma | 5 | 1 |
| Gastrointestinal carcinoma | 4 | 0,8 |
| Hepatocellular carcinoma | 2 | 0,4 |
| Thyroid carcinoma | 2 | 0,4 |
| Ovarian sex cord stromal tumour | 4 | 0,8 |
| Germ cell tumour | 3 | 0,6 |
| Neuro-endocrine tumour | 2 | 0,4 |
| Serous borderline tumour | 1 | 0,2 |
| Skin cancer | 1 | 0,2 |

### Germline findings in study cohort

A total of 130 (26.9%) patients had ≥1 GF (total n= 149) involving 74 genes (132 PGVs, 8 low penetrant/risk-alleles, 3 VUS+), one chromosomal condition (T21, n=5) and one molecularly-negative clinical diagnosis of Beckwith-Wiedemann syndrome (cBWS) with a score of 5 according to Brioude et al (10) (figures 2-3, table S2C). A total of 50 DFs were identified in 49/484 patients (10.1%), including one with two DFs; ten were diagnosed with a CPS prior to their primary cancer (figure 4). In 42/49 patients (85.7%), the DF was well*-*established: 28 had monoallelic PGVs in autosomal dominant CPGs, 5 had T21 (including one mosaic), 4 had other mosaic PGVs, 3 had bi-allelic PGVs in autosomal recessive CPGs, one had a monoallelic *ATM* PGV with a clear A-T phenotype, and one had cBWS. Together, these established DFs span over 21 genes (6% of the full gene panel), the most frequent being *TP53* (n=7), *RB1* (n=4), *SMARCB1* (n=3). Eight DFs were classified as suspicious, due to the patient’s phenotype (figure 2): two were non-typical cancer-gene associations (*DICER1* PGV in neuroblastoma and *PTPN11* PGV in T-ALL), two were monoallelic *PMS2* PGVs in mismatch repair-deficient (MMRd) B-ALL and AML; and one consisted of a *DDX41* PGV, usually associated with adult-onset AML, in an infant with mosaic T21 who developed AML. The three remaining patients harbored VUS+ in genes (*TP53*, *RECQL4* and *FH*) that were well associated with their cancer phenotypes (Supplementary Results).

**Figure 2.**
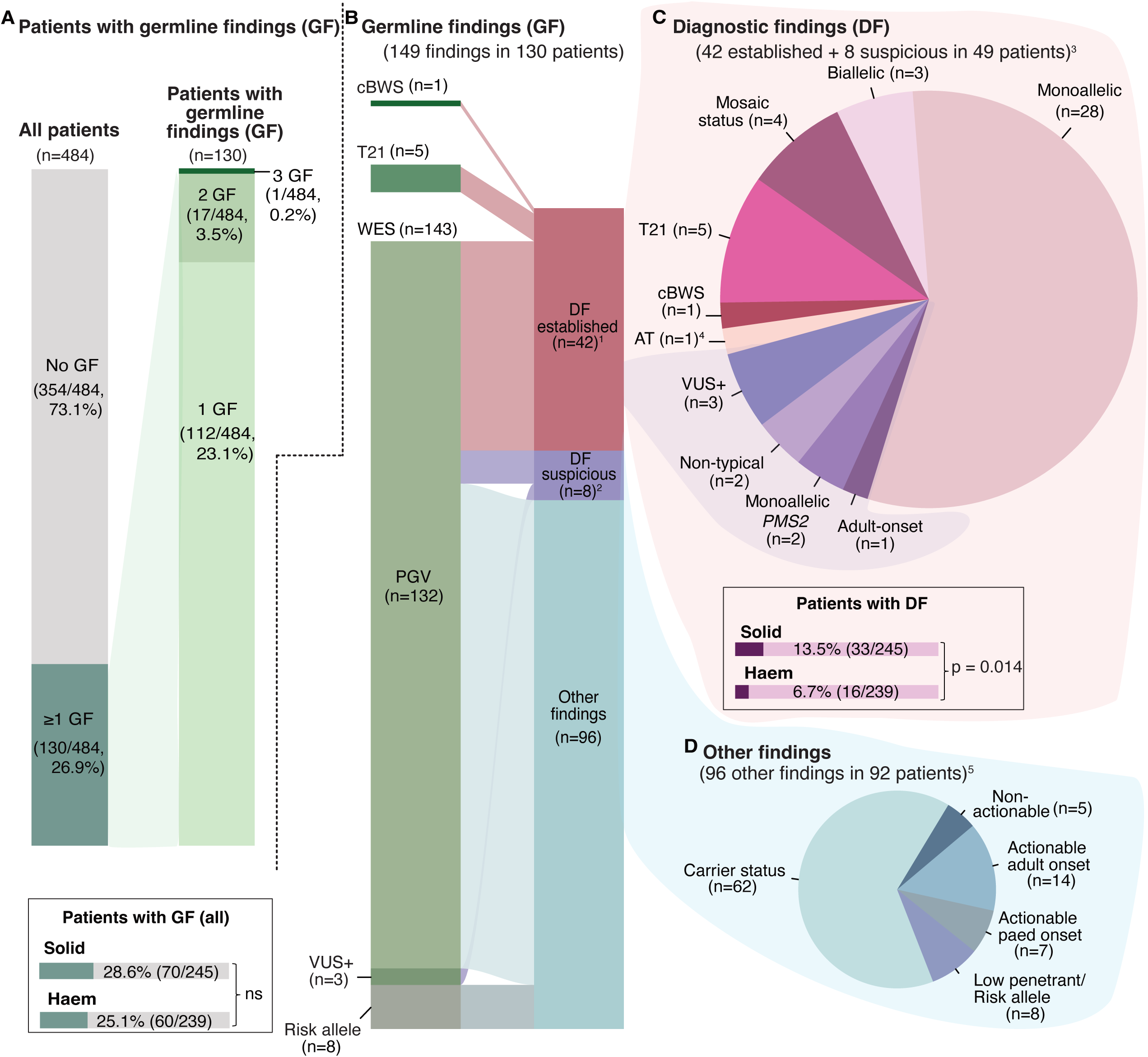
Prevalence and distribution of germline findings in the Quebec paediatric cancer cohort. A. Proportion of patients with ≥1 germline finding (GF) in the entire cohort (n=484) and number of GF per patient among patients with ≥1 GF (130/484, 26.9%). B. Classification of germline finding (n=149), including WES-based findings, trisomy 21, and one patient with clinical diagnosis of Beckwith–Wiedemann syndrome (cBWS). The alluvium is colored according to association subgroups. C. Subcategories of diagnostic findings (DFs) in 49 patients (one with both an established and a suspicious DF). Established DFs (n=42): monoallelic; biallelic; mosaic status; T21; cBWS; and monoallelic *ATM* pathogenic/likely pathogenic germline variant (PGV) with ataxia-telangiectasia phenotype (AT). Suspicious DFs (n=8): highly suspicious variant of uncertain significance (VUS+); non-classical tumour presentations; monoallelic *PMS2* PGV with mismatch repair deficiency; and PGV associated with adult-onset cancer predisposition syndrome but typical tumour. D. Subcategories of other findings (OF; n=96): actionable-paediatric onset; actionable-adult onset; non-actionable; carrier status; low penetrance/risk allele. Notes: 1: Established DFs include two biallelic *BRCA2* PGVs consistent with Fanconi anemia (FA-D1). 2: Suspicious DFs include one biallelic *RECQL4* state (one PGV + one VUS+), consistent with probable Rothmund–Thomson syndrome. 3: One infant carried both mosaic T21 (established DF) and a *DDX41* PGV (suspicious DF). 4: Patient with monoallelic *ATM* PGV and AT. 5: One patient had three other findings (all carrier status); two patients had two other findings each (paediatric-onset CPS + adult-onset CPS; and risk allele + adult-onset CPS). **Abbreviations:** AR, autosomal recessive; AT, ataxia-telangiectasia phenotype; cBWS, clinical Beckwith–Wiedemann syndrome; DF, diagnostic finding; GF, germline finding; Haem, haematologic neoplasms; Htz, heterozygous; ns, non-significant; OF, other finding; PGV, pathogenic/likely pathogenic germline variant; p, p value; Solid, solid cancers; paed, paediatric; T21, trisomy 21; VUS+, highly suspicious variant of uncertain significance.

**Figure 3.**
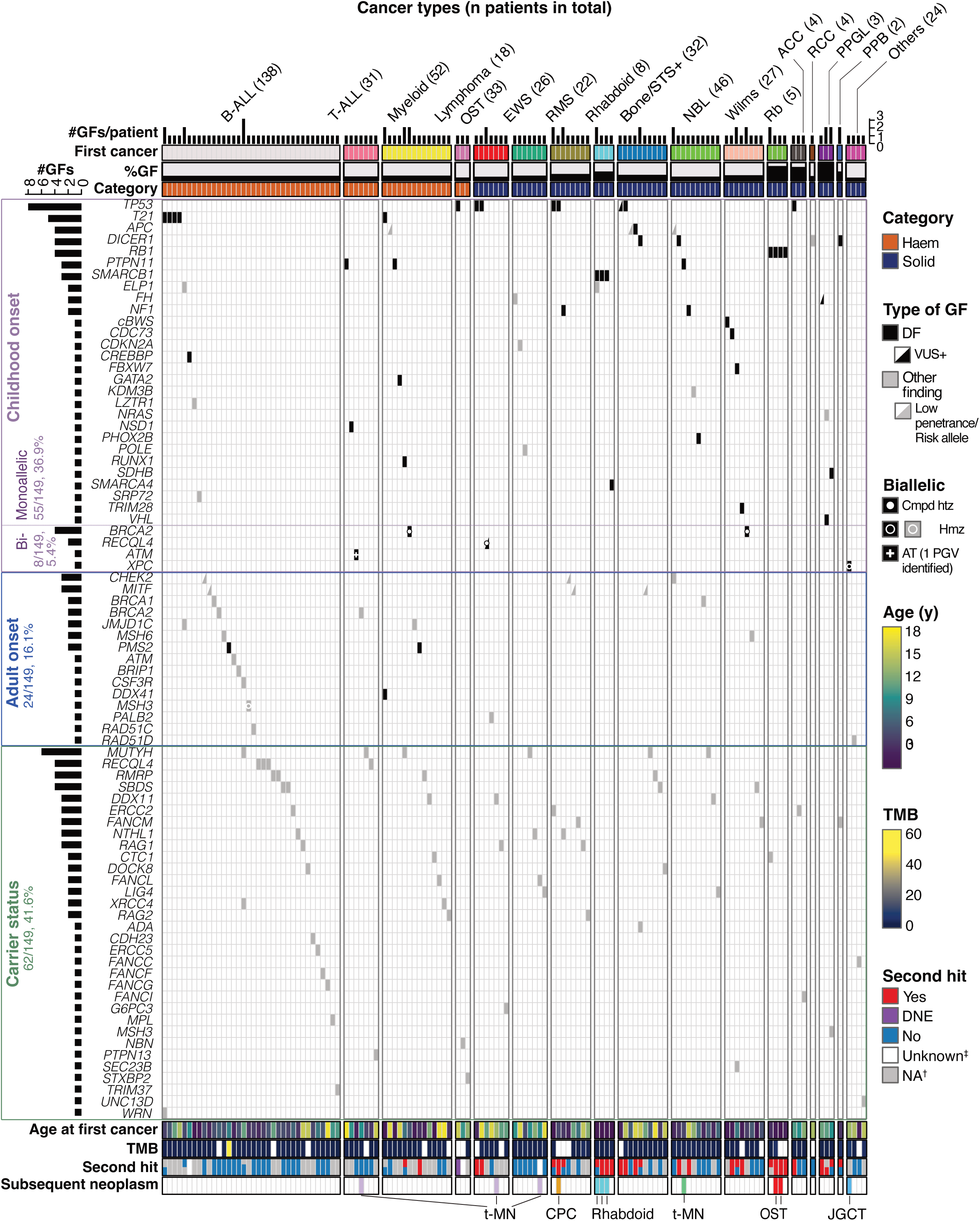
Landscape of germline findings in paediatric cancer patients. Oncoprint representation restricted to patients harboring ≥1 germline finding (GF; n=130, 26.9% of the cohort), stratified by cancer type and by gene-level category of cancer predisposition (paediatric-onset, adult-onset, or carrier status). Each column represents one patient, and each row corresponds to a gene or recurrent syndrome. Cancer types are grouped into haematologic and solid categories and sorted by decreasing frequency within the cohort^1^. Numbers in parentheses above each tumour type indicate the total number of patients with that cancer in the entire cohort (not limited to those with a GF). The barplot above the heatmap indicates the number of GFs per patient, and the barplot on the left of the heatmap indicates the number of GFs in each gene or syndrome (compound heterozygous variants resulting in a double count for one patient). Additional upper annotations include the first cancer type, proportion of GF carriers within each cancer type (%GF), category of cancer. Lower annotations encompass age at first cancer diagnosis, tumour mutational burden (TMB), second-hit status for monoallelic variants in tumour suppressor genes (TSG) (displayed in the same order as in the heatmap when multiple GFs are present and not applicable for trisomy 21, biallelic syndromes, or non-clear TSGs), and the occurrence of subsequent neoplasms. Categorization of each variant as diagnostic finding (DF, black cells) or other finding (OF, grey cells), as well as variant-specific information, is shown directly in the heatmap. **Symbols:** ‡, tumour whole-exome sequencing data unavailable; †, second hit not applicable (T21, biallelic syndrome, or monoallelic PGV in non-clear TSG gene) **Notes:** 1: Hepatoblastomas are not displayed, as no patient carried a GF **Abbreviations:** AT, ataxia-telangiectasia; ACC, adrenocortical carcinoma; B-ALL, B-cell acute lymphoblastic leukemia; Bone/STS+, other bone or soft-tissue sarcomas; Cmpd htz, compound heterozygous; CPC, choroid plexus carcinoma; DF, diagnostic finding; DNE, PGV with dominant-negative effect; EWS, Ewing sarcoma; GF, germline finding; Haem, haematologic neoplasm; Hmz, homozygous; JGCT, juvenile germ cell tumour; NA, not applicable; NBL, neuroblastoma; OF, other finding; OST, osteosarcoma; PGV, pathogenic or likely pathogenic germline variant; PPB, pleuropulmonary blastoma; PPGL, paraganglioma/pheochromocytoma; Rb, retinoblastoma; RCC, renal cell carcinoma; RMS, rhabdomyosarcoma; Solid, solid cancers; T−ALL, T-cell acute lymphoblastic leukemia; TMB, tumour mutational burden; t-MN, therapy-related myeloid neoplasm, TSG, tumour suppressor gene; VUS+, highly suspicious variant of uncertain significance; y, years.

**Figure 4.**
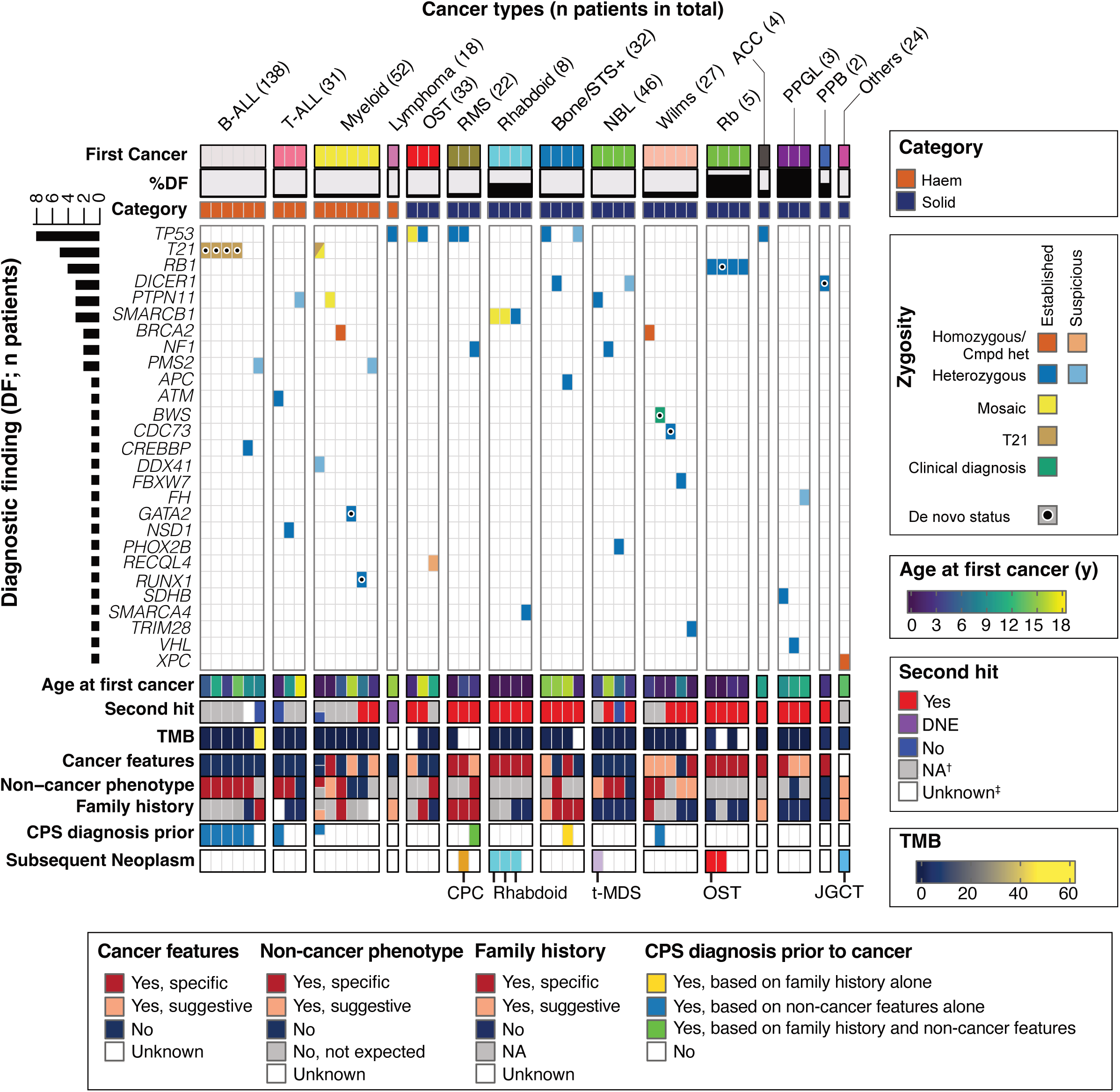
Heatmap of diagnostic findings across paediatric cancer patients. Oncoprint representation restricted to patients harboring ≥1 diagnostic finding (DF; n=49, 10.1% of the cohort), stratified by cancer type and by cancer predisposition syndrome. Each column represents one patient, and each row corresponds to a gene or recurrent syndrome. Cancer types are grouped into haematologic and solid categories and sorted by decreasing frequency within the cohort1. Numbers in parentheses above each tumour type indicate the total number of patients with that cancer in the entire cohort (not limited to those with a DF). The barplot on the left of the heatmap indicates the number of DFs in each gene or syndrome (compound heterozygous variants resulting in a unique count for one syndrome in one patient). Upper annotations include first cancer type, proportion of DF carriers within each cancer type (%DF), category of cancer (haematologic or solid). Lower annotations encompass age at first cancer diagnosis; second-hit status for monoallelic variants in clear tumour suppressor genes (TSG) (displayed in the same order as in the heatmap when multiple DFs are present, and considered not applicable for trisomy 21, biallelic syndromes, and non-clear TSGs); tumour mutational burden (TMB); cancer features; non-cancer phenotype and their association with the underlying CPS; family history; diagnosis prior to or after cancer and, if yes, reason for diagnosis; occurrence of subsequent neoplasms. **Symbols:** ‡, tumour whole-exome sequencing data unavailable; †, second hit not applicable (T21, biallelic syndrome, or monoallelic PGV in non-clear TSG gene). Notes: 1: Hepatoblastomas, and Ewing sarcomas are not displayed, as no patient with this cancer type carried a DF **Abbreviations:** B-ALL, B-cell acute lymphoblastic leukemia; Bone/STS+, other bone or soft-tissue sarcomas; BWS, Beckwith–Wiedemann syndrome; Cmpd htz, compound heterozygous; CPC, choroid plexus carcinoma; CPS, cancer predisposition syndrome; DF, diagnostic findings; DNE, PGV with dominant-negative effect; Haem, haematologic neoplasms; JGCT, juvenile germ cell tumour; NA, not applicable; NBL, neuroblastoma; OST, osteosarcoma; PPB, pleuropulmonary blastoma; PPGL, paraganglioma/pheochromocytoma; Rb, retinoblastoma; RCC, renal cell carcinoma; RMS, rhabdomyosarcoma; Solid, solid cancers; STS, soft tissue sarcoma; T21, trisomy 21; T−ALL, T-cell acute lymphoblastic leukemia; TMB, tumour mutational burden; t-MDS, therapy-related myelodysplastic syndrome; TSG, tumour suppressor gene; y, years.

A total of 96 other findings were present in 92 patients (19%), including 11 who also had a DF. Twenty-one (4.3%) were actionable, seven of which (1.4%) were clinically actionable in childhood. These seven PGVs were identified in children with tumour types unrelated to the affected gene and without somatic evidence supporting causality (e.g., second hit, pathway alteration, immunohistochemistry) (table S3) (Supplementary Results). Fourteen (2.9%) patients had an actionable other finding for an adult-onset CPS (monoallelic n=12, biallelic n=1) (table S3), with half included in the ACMG v.3.3 list of reporting of secondary findings (11): *BRCA1* (n=2), *BRCA2* (n=2), *MSH6* (n=2), and *PALB2* (n=1). A monoallelic PGV for an autosomal recessive CPS was identified in 61 (12.6%) patients. Additionally, 8 patients (1.7%) had reduced-penetrance/risk alleles for adult-onset cancers (table S2C; Supplementary Results). Amongst the full cohort, five PGVs with likely no actionability were detected in four patients, one of whom also had a DF (*SMARCB1* and *ELP1* PGVs in a patient with a rhabdoid tumour).

### Frequency and relevance of GFs across cancer types and clinical trajectories

DFs were more frequent in patients with solid tumours than in patients with hematological malignancies (13.5% [33/245] vs 6.7% [16/239] respectively, p=0.014). Frequencies of DFs and number of genes/conditions involved varied markedly by cancer type. Highest rates were seen in certain rare malignancies classically known to be associated with CPSs, including pheochromocytoma and paraganglioma (PPGL), pleuropulmonary blastoma (PPB), retinoblastoma, and rhabdoid tumours (all had DF rates of ≥50%). All DFs identified in the aforementioned tumours spanned over 1-3 genes per cancer type (figure 5). When looking at the most common cancer types in the cohort (n ≥20), frequencies of GFs steadily ranged from 21.2-36.4% while frequencies of DFs varied between 0% (EWS) to 18.5% (Wilms). Among 15 patients with known multiple primary cancers (subsequent malignant neoplasms n=14, synchronous cancers n=1), 11 (73%) had a GF (8 with DFs, 3 with other findings) (figure 3). DFs were identified in *SMARCB1* (n=3), *RB1* (n=2), *TP53* (n=1, *XPC* (n=1, biallelic), and *PTPN11* (n=1) (figure 4). Six patients had therapy-related myeloid neoplasms, one with a PGV in *PTPN11* and three with monoallelic PGVs in each of *BRCA2*, *DDX11*, *FANCL*. The contribution of such PGVs involved DNA-repair pathways in the development of therapy-related myeloid neoplasms merits further investigation.

**Figure 5.**
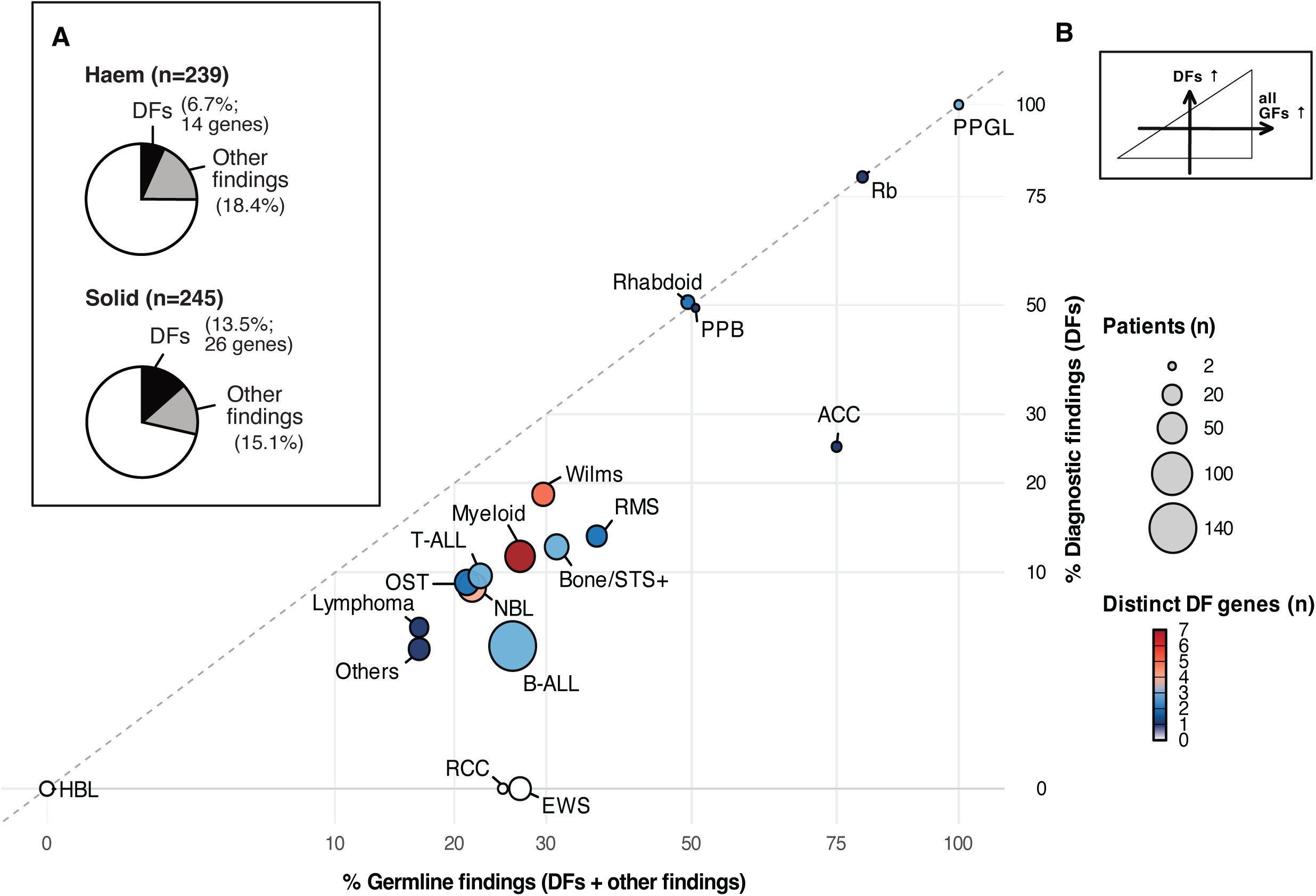
Diagnostic yield across cancer types. A. Proportion of diagnostic findings (DFs) within haematologic and solid cancers. The black segments indicate the proportion of DFs, whereas the grey segments indicate the proportion of other findings. When both a DF and other finding(s) are identified in the same patient, the DF overrides other findings. B. Scatterplot illustrating, for each cancer type, the proportion of patients with any germline finding (DF+other finding, x axis) versus the proportion carrying a DF (y axis). Both axes are displayed on a square-root scale. Each dot represents a cancer type; color indicates the number of distinct genes classified as DF in that cancer type (gradient from dark blue [1 gene] to dark red [7 genes]; white indicates no DF), and circle area is proportional to the number of patients. The dashed diagonal line represents parity between overall germline findings detection and clinically relevant findings (DF). **Abbreviations:** ACC, adrenocortical carcinoma; B-ALL, B-cell acute lymphoblastic leukemia; Bone/STS+, other bone or soft-tissue sarcomas; DF, diagnostic findings; EWS, Ewing sarcoma; GF, germline finding; Haem, haematologic neoplasms; HBL, hepatoblastoma; Myeloid, myeloid malignancies; NBL, neuroblastoma; OF, other findings; OST, osteosarcoma; Others, other malignancies; PPB, pleuropulmonary blastoma; PPGL, pheochromocytoma/paraganglioma; Rb, retinoblastoma; RCC, renal cell carcinoma; RMS, rhabdomyosarcoma; Solid, solid cancers; T-ALL, T-cell acute lymphoblastic leukemia.

### Integrated Somatic Data Informs Germline Variant Interpretation

Matched somatic data from tumour WES and RNA sequencing provided valuable insights for interpreting germline variants.

#### Somatic second hits

Among 34 patients with DF involving monoallelic variants in autosomal dominant tumour suppressor gene, somatic WES was available for 33, with a second hit identified in 29/33 cases (87.9%), including 25 established DF (figure 4, table S4). LOH was observed in four suspicious DFs (*DICER1* PGV in neuroblastoma, *PMS2* PGV in AML, *FH* VUS+ in RCC, *TP53* VUS+ in infantile fibrosarcoma), whereas no second hit was identified among 57 other findings in the available somatic data (p-value <0.0001) (figure 3).

#### Mosaic

Detecting mosaic GFs can be challenging because of their low allelic frequencies (8–21% in this cohort, corresponding to only 6–30 sequencing reads) combined with typically lower germline testing coverage. All four mosaic PGVs were confirmed by heterozygous or homozygous allelic frequencies in somatic WES (figure 6A), highlighting its utility in uncovering mosaic GFs that might otherwise be missed.

**Figure 6.**
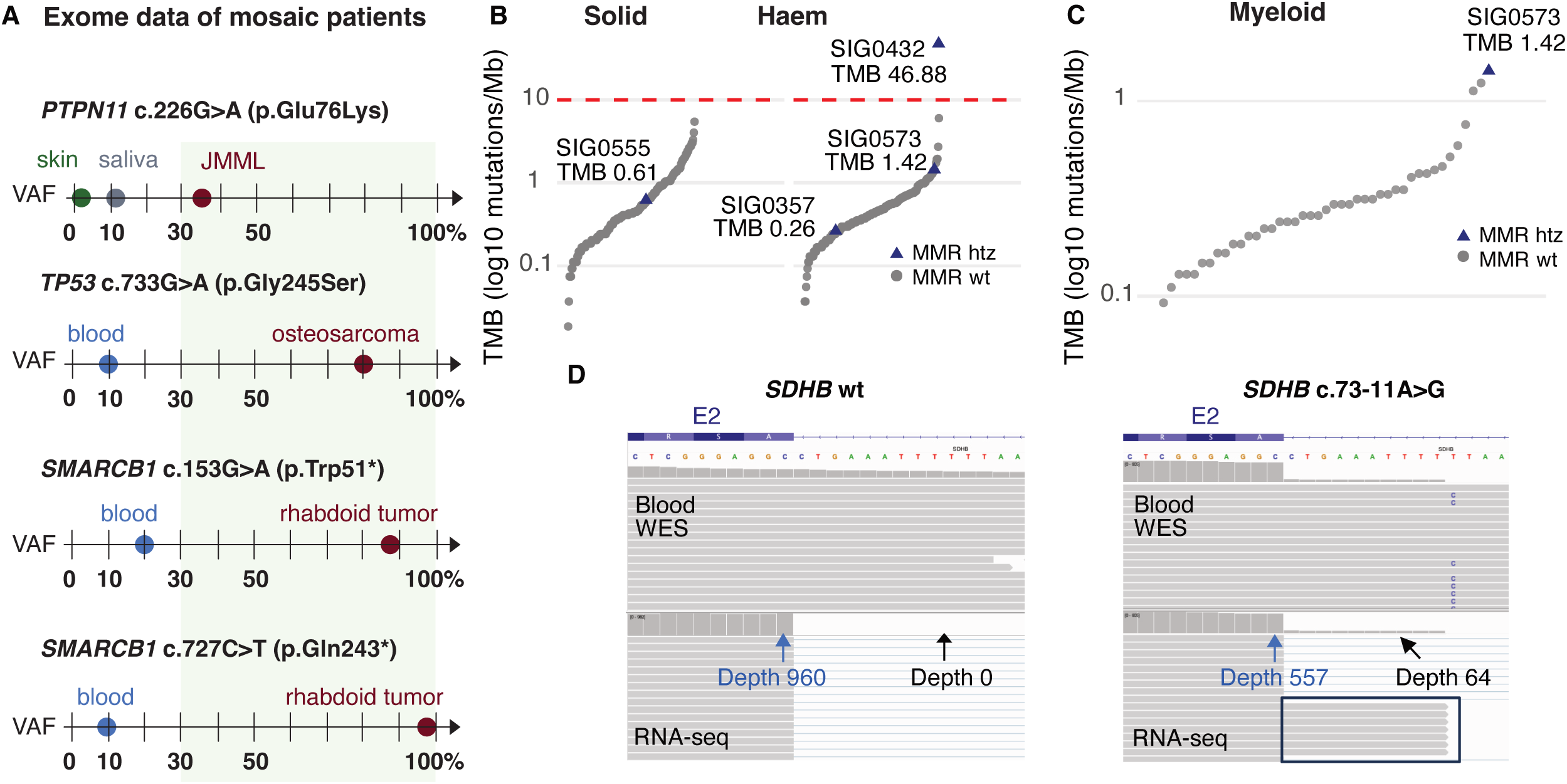
Integrated somatic data for assessment of germline variant pathogenicity. A. Exome data from 4 mosaic patients. Variant allele frequencies (VAFs) in germline DNA (skin, saliva, and/or blood) were consistently below 30%, which was the filtering threshold for germline variant calling (AF ≥ 0.3), and would therefore have been missed by standard germline pipeline. Somatic sequencing was essential for variant detection. Comparison with raw germline data confirmed mosaicism and excluded sequencing artefacts. B. Tumour mutation burden (TMB) across the cohort, displayed on a log10-scaled y-axis, with tick marks indicating absolute TMB values (mutations per Mb). The red dashed line indicates the hypermutation threshold (10 mutations/Mb). Patients carrying a heterozygous pathogenic or likely pathogenic variant (PGV) in MMR genes are shown as blue triangles and labelled with patient identifiers. SIG0432 is hypermutated (TMB > 10 mutations/Mb) and shows the highest TMB in the entire cohort. MMR htz: patients with a heterozygous PGV in a MMR gene; MMR wt: patients without PGV in MMR genes. C. TMB distribution among myeloid neoplasms (n=43), highlighting SIG0573 as the case with the highest TMB within this subgroup. Notably, this patient is the only with a heterozygous MMR PGV among myeloid malignancies. D. Integrative Genomics Viewer (IGV) visualization of whole exome sequencing (WES) and transcriptome data at the *SDHB* exon 2 locus (transcript NM_003000.3), comparing a heterozygous carrier of the *SDHB* c.73-11A>G variant (right) to a wild type individual (left). In the carrier, germline WES data (upper track) confirm the heterozygosity (asterisk), with the variant located 11 bp upstream of exon 2. Transcriptome data (lower track) demonstrate retention of 10 nucleotides from intron 1 at the 5’ end of exon 2 (black box), consistent with activation of a cryptic acceptor splice site. In contrast, the control sample shows no variant at the DNA level and displays canonical exon-exon junctions without intronic retention. Read-depth values are indicated at key positions. **Abbreviations:** AF, allele frequency; JMML, juvenile myelomonocytic leukemia; Haem, haematologic neoplasms; MMR, mismatch repair; RNA-seq, RNA sequencing data; Solid, solid cancers; TMB, tumour mutation burden; VAFs, variant allele frequencies; WES, whole exome sequencing.

#### Tumour Mutational Burden (TMB)

TMB and mutational signatures were performed in 377 (78%) and 224 (46%) patients respectively (figure 6B). Two patients with leukemia with monoallelic *PMS2* PGVs showed evidence of MMR dysfunction: (1) SIG0432 with B-ALL had the highest TMB in the cohort (47 mutations/Mb) and MMR-related signatures (SBS26, SBS20), despite no second hit detected (figure 5B); the *PMS2* variant was paternally inherited, and maternal history suggested Lynch syndrome, raising the possibility of an undetected alteration. (2) SIG0573 with AML, had the highest TMB among myeloid neoplasms (1.4 mutations/Mb), *PMS2* LOH, and an MMR signature (SBS15, figure 5C). Two additional patients with *MSH6* PGVs (B-ALL, bilateral Wilms tumour) showed low TMB and no MMR signatures and were classified as other findings. Immunohistochemistry was not performed in either case. No solid tumours exhibited profiles suggestive of MMR impairment.

#### Splicing

Eighteen GFs (including 3 DFs) affected canonical splice sites and splice regions (table S5). Tumour RNA sequencing confirmed aberrant splicing defects and provided functional evidence supporting variant interpretation. In one sample, RNA sequencing revealed a 10bp intronic retention in *SDHB* (c.73-11A>G) in a child with metastatic paraganglioma, leading to the upgrade of the intronic VUS to a LP variant (figure 6D).

### Phenotype-genotype correlations

Of the 49 patients with a DF, 10 (20%) had a CPS diagnosed prior to first cancer development (figure 4). CPSs were diagnosed based on clear non-cancer phenotypic features alone (n=8), family history alone (n=1), or both (n=1) (figure 4, table S6). In the 39 patients where the CPS diagnosis was subsequent to their cancer presentation, 36 (92.3%) had one or more suggestive and/or specific features in their cancer presentation (n=23), non-cancer phenotypes (n=12) and/or family history (n=12), raising suspicion for CPS. In 25/39 patients, these features pointed towards specific CPSs, including retinoblastoma in patients with *RB1* PGVs and a *TP53* PGV in a young-onset anaplastic rhabdomyosarcoma with a family history meeting Chompret criteria. Cancer presentation alone was the sole suggestive and/or specific feature raising suspicion for CPS in 16/39 children. Non-cancer phenotypes were specific for related CPSs in 8/18 (44.4%) patients (e.g, Fanconi anemia with hyperpigmented macules and extra digit, Rothmund-Thomson syndrome with classic growth and cutaneous features). Family history of cancer was the sole feature raising suspicion for CPS in 4/38 (10.5%) patients with available family history information (and without a known CPS at the time of cancer presentation). Three (7.5%) patients had no suspicious features for an underlying CPS: AML with a *RUNX1* PGV (SIG0625), neuroblastoma with a *DICER1* PGV (SIG0323), and T-ALL with a *PTPN11* PGV (TRI0230).

Seven PGVs, including six DFs, were initially missed by standard germline exome pipelines due to technical limitations (table S3). Six involved microdeletions detected by MLPA or microarray (*RB1* n=2, *BRCA2* n=2, *CREBBP*, *SMARCB1)*, following phenotype-driven reanalysis. For example, a patient with bilateral Wilms tumour and positive chromosomal breakage had a single *BRCA2* PGV identified through standard pipelines, while a second alteration, a mono-exonic deletion involving *BRCA2*, was detected by microarray. The remaining PGV was an 8 bp duplication in *PHOX2B* (NM_003924.4:c.691_698dup) located in a GC-rich region with poor coverage (depth=9), recovered on a reanalysis prompted by the association of neuroblastoma and Hirschsprung disease. These findings underscore the importance of complementary strategies when clinical suspicion of a CPS is high.

## DISCUSSION

Reports describing GFs and cancer predisposition in paediatric cancer datasets exhibit recurring challenges. Variability in cohorts, reporting approaches, data comprehensiveness, and terminologies results in wide ranges of GF prevalence, and limits cross-study comparability. Moreover, current variant classification frameworks, which rely on categorical distinctions (e.g., PGV vs VUS), fail to capture the biological complexity observed in clinical practice. This study aimed to address these issues by characterizing the germline mutational portrait and integrating it with phenotypic, pathology, and molecular tumour features in 484 children enrolled in a province-wide paediatric oncology research network in Quebec. Our results underscore both the diagnostic yield and the required nuance in the interpretation of germline DNA sequencing in paediatric oncology.

GFs were identified in 130 (26.9%) of children with primary non-CNS malignancies, with 49 (37.7%) having a CPS with an established and/or suspicious association to their cancer, corresponding to a global CPS frequency of 10.3% in our cohort. Frequency of DFs varied according to malignancy type, with the highest rates seen in certain rare (and expected) tumours (e.g, retinoblastoma, PPGL) and much lower rates in some of the more common cancers (B/T-ALL, neuroblastoma, EWS and osteosarcoma).

Two percent (10/484) of children had a CPS diagnosis prior to cancer development. Aligned with what is expected, these CPS conditions are associated with particularly recognisable non-cancer phenotypes or highly penetrant familial conditions. In 80% of patients with a CPS, cancer was the first recognised manifestation. At the time of first cancer presentation, 92.3% of these children had features hinting towards a CPS, 69.4% of whom presented at least one CPS-specific feature. This highlights the importance of comprehensive history taking and clinical examinations at cancer diagnosis, with particular attention to growth parameters, distinctive facial features, and skin hyper-/hypopigmentation. This also underscores the need for oncologists to remain updated on cancer types and related features associated with CPS (12). Family history was the sole feature raising suspicion for a hereditary condition in only 10% of children with a CPS, reflecting the frequency of *de novo* events, incomplete penetrance of CPSs, and the recessive nature of some CPSs amongst other factors (2). Three children with actionable paediatric-onset CPSs were identified only through germline sequencing and had no suspicious phenotypes. Conversely, phenotypic information aided in identifying CPSs without (cBWS) or with incomplete (A-T and monoallelic *ATM* PGV) molecular confirmation. Rather than being mutually exclusive, phenotypic and genomic-based approaches provide complementary insights that enhance diagnostic precision and clinical relevance.

Germline DNA sequencing using WES presents limitations that are important to recognise. Next-generation short read sequencing approaches are less performant for detecting alterations in repetitive or low-complexity regions and in the presence of pseudogenes. WES also shows reduced sensitivity for small CNVs. These limitations were seen in seven children in this cohort who required alternative approaches to confirm diagnoses. Furthermore, standard WES protocols typically aim for lower sequencing coverage, which can fail to detect low-level mosaic variants, underscoring the added diagnostic yield of tumour analysis in revealing clinically relevant mosaicism. Nevertheless, an advantage of unbiased genomics such as WES is its intrinsic flexibility in an evolving field, enabling adaptive gene panels and opportunity for re-analysis; in our cohort, recently discovered PGVs that would likely have been missed using fixed targeted panels were identified (e.g., *FBXW7*). In a research context where mandates of germline investigations extend beyond aspects of causality and actionability, an unselected germline sequencing approach appears optimal when feasible and with appropriate pre-and post-test counselling (Bakhuizen 2024). Paired germline–somatic sequencing decreases the risk of misreporting germline variants as tumour-specific alterations in addition to streamlining somatic analyses.

Practical challenges directly impacting clinical practices emerged from this initiative. First, we identified GFs clinically actionable in childhood, unrelated to the individual’s cancer. Clinicians must manage uncertainties in causality, consider emerging associations, and ensure appropriate surveillance and follow-up for these incidental CPSs. Secondly, adult-onset CPSs with potential timely actionability for relatives, identified in 3% of our cohort, pose a challenge. These GFs are not typical “secondary findings”, as they are on the panel due to their implication in biallelic settings (e.g., *ATM*, *BRCA2*, *PALB2*, MMR-related genes). Accordingly, pre-test counseling and consent discussions should explicitly address these scenarios, rather than relying on standard secondary-finding frameworks. Furthermore, PGVs in genes typically associated with adult-onset cancers may contribute to paediatric tumourigenesis under specific genetic backgrounds, challenging current dichotomies between childhood-and adult-onset predisposition. In our cohort, two monoallelic *PMS2* PGVs were considered suspicious DFs with the integration of somatic evidence (TMB, LOH, MMR mutational signatures). More broadly, monoallelic PGVs in DNA-repair genes increasingly raise oncologists’ concerns around treatment-related toxicity (13–15). While causality may be uncertain, clinicians must still integrate these findings into therapeutic decision-making. In addition, all patients with a CPS should be offered genetic counseling and cascade testing in at-risk family members. Finally, the use of a virtual panel leads to the identification of PGVs in genes with likely no actionability (1.0%), risk alleles (1.7%), and heterozygous variants for autosomal recessive disorders (12.6%). These findings are not rare and there remains inconsistent guidance on how to manage them.

Study limitations are acknowledged. First, although this cohort does not represent a populational study, the gradual extension of eligibility and improved coordination of precision oncology initiatives have led to steadily increasing participation rates (>90%) in Quebec. Secondly, given the nature of retrospective data collection, it is expected that quality and comprehensiveness of clinical information differ between patients. Information on subsequent malignancies was not ascertained in some participants due to lack of follow-up information. As such, we avoided comparative analyses between participants with/without GFs or specific outcomes, as the clinical documentation was suboptimal in participants without GFs. Despite these limitations, our provincial research experience reveals the analytical and practical considerations of germline evaluations and supports a shift toward integrative interpretation models that combine germline, somatic, pathological, cytogenetic, and phenotypic data, rather than relying predominantly on variant classification frameworks. Multidisciplinary case discussions combining clinical, laboratory and research expertise are essential to ensure rigorous and context-sensitive interpretation. Whether clinical or research-based precision oncology initiatives, ethical challenges arise in all steps of investigations and communications, most often during pre-test counseling and patient/caregiver consent process. Similarly, documentation of clinical features and family histories in a pre-test manner can facilitate variant interpretation and reporting queries.

## CONCLUSION

This study provides a comprehensive view of germline predisposition across paediatric malignancies, reflecting nearly a decade of integration of genomic medicine in paediatric oncology practice in the province of Quebec. This research effort has been a pillar in justifying the implementation and navigation of a publicly-funded mainstreaming approach in paediatric oncology. This combined research and clinical program now serves approximately 300 children with cancer annually and leverages the multidimensional and multi-institutional expertise. The expansion of collaborative precision oncology initiatives and the evolution of sequencing technologies will improve our collective ability to refine risk assessment, to optimize patient management, and to translate genomic discoveries into tangible benefits for affected children and their families.

## Ethical considerations

The study was approved by the CHU Sainte-Justine Research Ethics Board (Triceps #2014-668; Signature #2019-2032). Written informed consent was obtained for all patients included in the present study.

## Supporting information

Supplementary Results

## Data Availability

The data generated in this study, including sequencing data, are not publicly available due to ethical and legal restrictions related to human genomic data. Aggregated data are provided in the manuscript and Supplementary Materials. Individual-level data may be made available upon reasonable request to the corresponding author, subject to institutional approval and applicable data access agreements.

## Acknowledgements and Funding

The authors thank the complete Signature team from all four participating institutions as well as the personnel at CRA-CHU Sainte-Justine biobank. The Signature project, including patient recruitment, biobanking, and data analysis, is generously funded by the Fondation Charles-Bruneau, with additional funds from the Fondation CHU Sainte-Justine and the Montreal Children’s Hospital Foundation. VPL, THT, RS and CG are supported by Clinician-Scientist Scholarships from the Fonds de recherche du Québec – Santé. VPL and CG received additional funding from the Cole Foundation. ES is supported by the CHU of Liège and by the Fonds Léon Frédéricq. This research was enabled in part by support provided by Calcul Québec (www.calculquebec.ca) and the Digital Research Alliance of Canada (alliancecan.ca).

## References

1. Kratz CP. Re-envisioning genetic predisposition to childhood and adolescent cancers. Nat Rev Cancer. 2025 Feb;25(2):109–28.

2. Zhang J, Walsh MF, Wu G, Edmonson MN, Gruber TA, Easton J, et al. Germline Mutations in Predisposition Genes in Pediatric Cancer. N Engl J Med. 2015 Dec 10;373(24):2336–46.

3. von Stedingk K, Stjernfelt KJ, Kvist A, Wahlström C, Kristoffersson U, Stenmark-Askmalm M, et al. Prevalence of germline pathogenic variants in 22 cancer susceptibility genes in Swedish pediatric cancer patients. Sci Rep. 2021 Dec 1;11(1).

4. Fiala EM, Jayakumaran G, Mauguen A, Kennedy JA, Bouvier N, Kemel Y, et al. Prospective pan-cancer germline testing using MSK-IMPACT informs clinical translation in 751 patients with pediatric solid tumors. Nat Cancer. 2021 Mar 1;2(3):357–65.

5. Khater F, Vairy S, Langlois S, Dumoucel S, Sontag T, St-Onge P, et al. Molecular Profiling of Hard-to-Treat Childhood and Adolescent Cancers. JAMA Netw Open. 2019 Apr 5;2(4):e192906.

6. Richards S, Aziz N, Bale S, Bick D, Das S, Gastier-Foster J, et al. Standards and guidelines for the interpretation of sequence variants: a joint consensus recommendation of the American College of Medical Genetics and Genomics and the Association for Molecular Pathology. Genet Med. 2015 May;17(5):405–24.

7. Clinical Genome Resource (ClinGen). ClinGen Variant Curation Expert Panels. 2026 [cited 2026 Feb 6]. Available from: https://clinicalgenome.org/affiliation/vcep

8. Schmidt RJ, Steeves M, Bayrak-Toydemir P, Benson KA, Coe BP, Conlin LK, et al. Recommendations for risk allele evidence curation, classification, and reporting from the ClinGen Low Penetrance/Risk Allele Working Group. Genet Med. 2024 Mar;26(3):101036.

9. Garrett A, Allen S, Durkie M, Burghel GJ, Robinson R, Callaway A, et al. Classification of variants of reduced penetrance in high-penetrance cancer susceptibility genes: Framework for genetics clinicians and clinical scientists by CanVIG-UK (Cancer Variant Interpretation Group-UK). Genet Med. 2025 Feb;27(2):101305.

10. Brioude F, Kalish JM, Mussa A, Foster AC, Bliek J, Ferrero GB, et al. Clinical and molecular diagnosis, screening and management of Beckwith-Wiedemann syndrome: An international consensus statement. Nat Rev Endocrinol. 2018 Apr 1;14(4):229–49.

11. Lee K, Abul-Husn NS, Amendola LM, Brothers KB, Chung WK, Gollob MH, et al. ACMG SF v3.3 list for reporting of secondary findings in clinical exome and genome sequencing: A policy statement of the American College of Medical Genetics and Genomics (ACMG). Genet Med. 2025 Aug;27(8):101454.

12. Goudie C, Witkowski L, Cullinan N, Reichman L, Schiller I, Tachdjian M, et al. Performance of the McGill Interactive Pediatric OncoGenetic Guidelines for Identifying Cancer Predisposition Syndromes. JAMA Oncol. 2021 Dec 1;7(12):1806–14.

13. Kratz CP, Smirnov D, Autry R, Jager N, Waszak SM, Großhennig A, et al. Heterozygous BRCA1 and BRCA2 and Mismatch Repair Gene Pathogenic Variants in Children and Adolescents with Cancer. J Natl Cancer Inst. 2022 Nov 1;114(11):1523–32.

14. Evans DG, Woodward ER. RE: Heterozygous BRCA1/BRCA2 and mismatch repair gene pathogenic variants in children and adolescents with cancer. J Natl Cancer Inst. 2023 Feb 8;115(2):224–5.

15. Daugs K, Brandes D, Yasin L, Anwar A, Alam J, Prasad Y, et al. Germline variants observed in pediatric cancer patients related to hereditary breast and ovarian cancer in adults. Int J Cancer. 2025 Dec 15;157(12):2447–54.

