## Supplementary figures and images for "Germline sequencing in children with cancer in Quebec: an integrated investigative approach"

### Figure S1

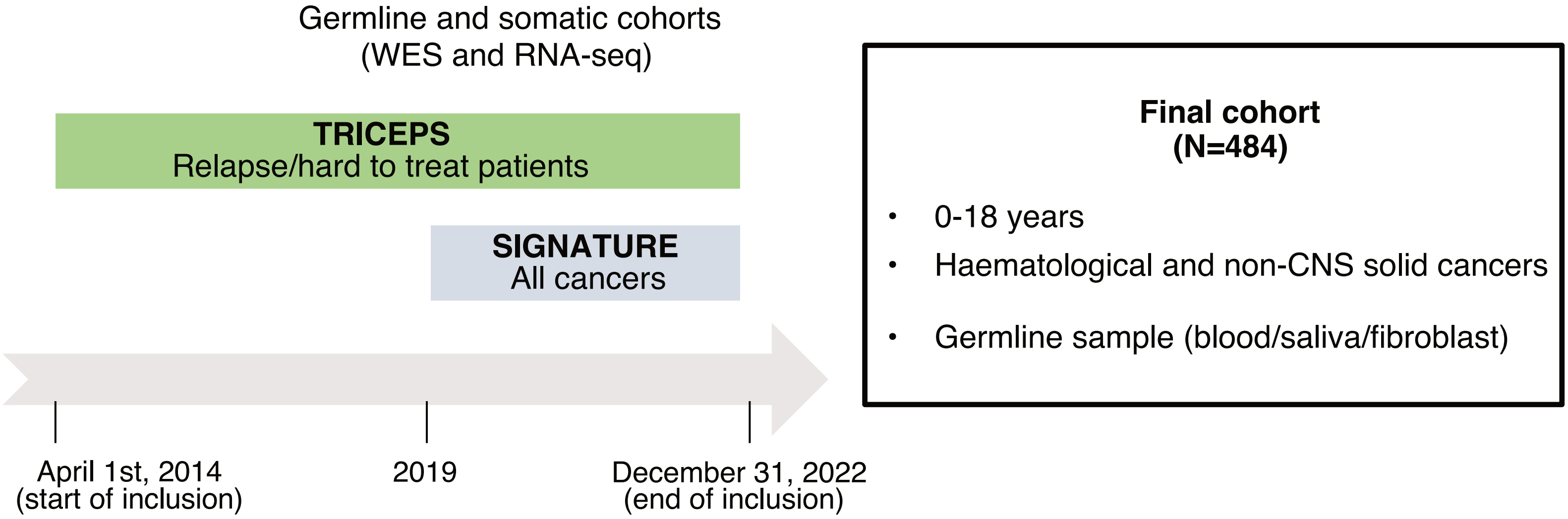

### Figure S2

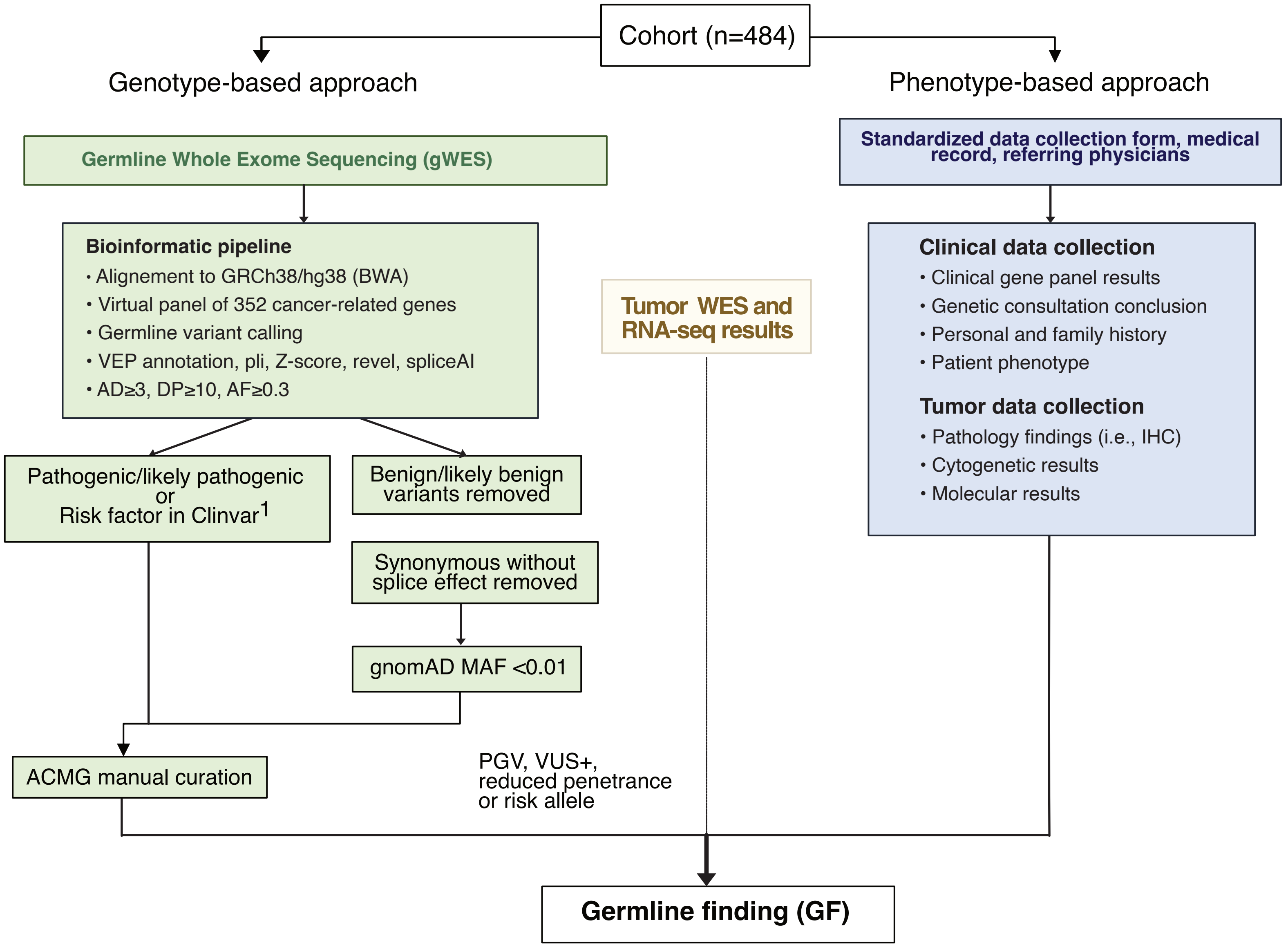
