## Supplementary material for "Germline sequencing in children with cancer in Quebec: an integrated investigative approach": Figure S2 legend

**Figure S2. Combined genotype- and phenotype-based approach.**

Overview of the genotype-based and phenotype-based approaches applied to the cohort (n=484). The genotype-based approach relied on a germline whole-exome sequencing (WES) analysed using a predefined 352-gene virtual panel and a standardised bioinformatic pipeline, including alignment, variant calling, annotation, filtering, and ACMG-based manual curation. Tumour WES and transcriptome were integrated when available. In parallel, the phenotype-based approach involved standardised collection of clinical and tumour data to contextualise germline findings and support final variant classification.

**Abbreviations:** ACMG, American College of Medical Genetics and Genomics; AD, allelic depth; AF, allele frequency; DP, read depth; IHC, immunohistochemistry; MAF, minor allele frequency; PGV, pathogenic or likely pathogenic variant; RNA-seq, RNA sequencing; VAF, variant allele frequency; VEP, Variant Effect Predictor; VUS+, highly suspicious variant of uncertain significance; WES, whole-exome sequencing.

Note: 1: Release 2022-11-29
