## Supplementary material for "Germline sequencing in children with cancer in Quebec: an integrated investigative approach": Figure S1 legend

**Figure S1. Enrolment timeline and genomic initiatives contributing to the final cohort.**

The final germline cohort (n=484) was assembled through two coordinated provincial sequencing initiatives in Quebec: TRICEPS, targeting relapse or hard-to-treat paediatric cancers, and SIGNATURE, including all paediatric cancers. Recruitment was conducted across the four paediatric haematology–oncology centres in Québec: CHU Sainte-Justine (Montreal), Montreal Children's Hospital (McGill University, Montreal), CHU de Québec–Université Laval (Quebec), and CHU Sherbrooke. Patients aged 0-18 years with haematological malignancies or non-central nervous system solid tumours and an available germline sample (blood, saliva, or fibroblasts) were eligible.

**Abbreviations:** CNS, central nervous system; RNA-seq, RNA sequencing; WES, whole-exome sequencing.
