## Supplementary Methods for "Germline sequencing in children with cancer in Quebec: an integrated investigative approach"

### DNA and RNA Sampling and Sequencing

Germline DNA was extracted from blood (solid tumours) or remission-phase blood or bone marrow, fibroblasts, or saliva (haematologic neoplasms).

Extractions were performed using Qiagen's AllPrep DNA/RNA Mini Kit or Oragene OG-250/500 kits (DNA Genotek). Exome capture was performed with the SureSelect XT Clinical Research Exome kit (Agilent Technologies), following the manufacturer's instructions. For transcriptomic analysis, total RNA previously extracted using the AllPrep DNA/RNA Mini Kit (Qiagen) was processed with the TruSeq Stranded Total RNA with Ribo-Zero Gold kit (Illumina), which includes ribosomal RNA depletion and library preparation.

Paired-end sequencing of matched germline and tumour DNA and stranded RNA libraries was performed on Illumina HiSeq 2500/4000 platforms (2×75 bp, before 2017) or NovaSeq 6000 systems (2×100 bp, from 2017 onward) at the Integrated Clinical Genomics Centre in Paediatrics (CHU Sainte-Justine, Montréal). The targeted mean coverage was 250–300× for tumour samples and 80–100× for germline samples.

### Exome Data Processing and Bioinformatic Pipeline

The bioinformatic pipeline followed GATK Best Practices for germline SNP and indel discovery in human exome sequencing. Raw sequencing reads in FASTQ format were aligned to the human reference genome build GRCh38/hg38 using the Burrows–Wheeler Aligner (BWA; v0.7.17) (1). BAM files from all sequencing lanes corresponding to the same library were merged with Picard (v2.20.6; http://broadinstitute.github.io/picard). To generate final BAM files, duplicate reads were marked, and base quality scores were recalibrated using the Genome Analysis Toolkit (GATK; v4.1.2) (2). Coverage metrics (mean depth and target coverage) were generated using DepthOfCoverage (GATK v.3.8).

Germline single-nucleotide variants (SNVs) and indels were identified with the GATK toolkit (v4.1.2). HaplotypeCaller was run on BAM files to produce gVCF files, with the "-G StandardAnnotation" parameter for allele-specific data.

Variants were filtered using the VariantFiltration command with GATK hard-filtering thresholds as follows: QD < 2.0, QUAL < 30.0, SOR > 4.0, FS > 60.0, MQ < 40.0,

MQRankSum < –12.5, ReadPosRankSum < –8.0 (SNVs) and QD < 2.0, QUAL < 30.0, FS > 200.0, ReadPosRankSum < –20.0 (indels).

Cohort-level VCFs were merged and annotated using the Ensembl Variant Effect Predictor (VEP; v107) on GRCh38. Annotation integrated data from gnomAD, ClinVar, and COSMIC, as well as predictive scores such as REVEL and SpliceAI. Annotated variants were filtered to retain only those located in a predefined panel of 352 cancer-related genes (table S1). Candidate germline variants were further filtered using read depth and allele frequency thresholds (AD ≥ 3, DP ≥ 10, AF ≥ 0·3). A dedicated analysis for putative mosaic variants applied the same depth thresholds but restricted calls to those with AF < 0·3. Candidate variants were reviewed manually in raw germline data whenever clinical or somatic features raised suspicion. Germline mosaicism, rather than sequencing artefact, was confirmed when the variant was also detectable in somatic data.

Transcriptome and WES somatic data were processed as previously described (3,4).

**Variant prefiltering and curation workflow**

Variants were retained if they met the following criteria (figure S2):

1. **Variants reviewed by expert panel**

Variants annotated as pathogenic or likely pathogenic by expert panels (“reviewed by expert panel” annotation in Clinvar) were directly retained and considered as such.

0. **Prefiltering based on Clinvar**

Remaining variants were retained if (a) classified as (likely) pathogenic, risk factor, or variant of uncertain significance (VUS) in Clinvar (release 2022-11-29) or (b) minor allele frequency (MAF) < 0.01 in gnomAD v2·1·1 and v3·1·2, not annotated as benign or likely benign in Clinvar (release 2022-11-29) and not synonymous variants without predicted splice effects.

0. **Manual curation**

- Prefiltered variants were classified following ACMG 2015 guidelines, complemented by ClinGen gene-specific specifications (where available) and selected adaptations from recent expert recommendations (table S2A–B).

- Variants were assigned to pathogenic, likely pathogenic, likely pathogenic-reduced penetrance, risk allele, or variant of unknown significance classifications.

- Variants of unknown significance (VUS) were retained only if supported by compelling functional, molecular, or clinical evidence suggesting pathogenic relevance.

**Integration of somatic data and pathology findings for evaluation of variant impact**

The molecular impact of prioritized variants was evaluated through integration of multi-omic and clinical data. Somatic WES data were analyzed for evidence of second hit (biallelic inactivation, loss of heterozygosity [LOH], or elevated tumour mutational burden [TMB]). Transcriptomic data were examined for aberrant expression or splicing effects. Pathological findings, including abnormal or absent protein expression by IHC and microsatellite instability (MSI) status, were incorporated into variant interpretation.

**References**


1. Li H, Durbin R. Fast and accurate short read alignment with Burrows-Wheeler transform. Bioinformatics. 2009 Jul 15;25(14):1754–60.
2. Van der Auwera GA, Carneiro MO, Hartl C, Poplin R, Del Angel G, Levy-Moonshine A, et al. From FastQ data to high confidence variant calls: the Genome Analysis Toolkit best practices pipeline. Curr Protoc Bioinformatics. 2013;43(1110):11.10.1-11.10.33.
3. Tran TH, Langlois S, Meloche C, Caron M, Saint-Onge P, Rouette A, et al. Whole-transcriptome analysis in acute lymphoblastic leukemia: a report from the DFCI ALL Consortium Protocol 16-001. Blood Adv. 2022 Feb 22;6(4):1329–41.
4. Khater F, Vairy S, Langlois S, Dumoucel S, Sontag T, St-Onge P, et al. Molecular Profiling of Hard-to-Treat Childhood and Adolescent Cancers. JAMA Netw Open. 2019 Apr 5;2(4):e192906.
