## Supplementary Results for "Germline sequencing in children with cancer in Quebec: an integrated investigative approach"

### Details for variants of unknown significance

Three GF involved *VUS* in canonical cancer predisposition genes with molecular or clinical features strongly suggestive of pathogenicity.

- ***TP53*** c.685T>C (p.Cys229Arg) was identified in a child with myofibrosarcoma. The tumour showed LOH at the *TP53* locus, suggesting biallelic inactivation. The variant causes a missense substitution of a highly conserved cysteine residue to arginine (Cys229Arg) within the DNA-binding domain. It has a very low population frequency (0.000007953 in gnomAD v2; absent from gnomAD v3; 0.000003101 in gnomAD v4; PM2_supporting). In silico predictions are inconclusive (BayesDel = 0·09; Align-GVGD = C0; BP4). Functional data available in the literature are inconsistent: Kato et al (1) reported partially retained transactivation activity in yeast (30%), and Zerdoumi et al (2) also suggested alteration of transcriptional activity; in contrast, Wasserman et al (3) and Kotler et al (4) indicated no loss of function (noLOF). Giacomelli et al (5) and Kawaguchi et al (6) did not analyse this variant. These discordant results preclude application of PS3 at any strength. The variant has been reported in paediatric tumours, including 3 adrenocortical carcinoma (3x0·5points) (3), a sarcoma (0.5 point, internal data) and in a woman who developed a breast cancer in her 30s (0.5 point) (7), resulting in a total of 2.5 points (PS4_moderate). According to ClinGen TP53 VCEP v2.3.0, the variant remains classified as a variant of unknown significance (VUS).
- ***FH* p.Pro363Arg**, very rare variant (one allele in gnomAD v2.1.1 and in v3.1.2; PM2_supporting), predicted deleterious (PP3_M), was found in a pheochromocytoma with somatic *LOH*, supporting its potential relevance to tumourigenesis.
- ***RECQL4* p.Leu787Pro**, extremely rare (one allele in gnomAD v2.1.1, absent in v3.1.2; PM2_supporting) and predicted deleterious (PP3), likely in trans with a PGV, was detected in a patient with features consistent with Rothmund–Thomson syndrome, a phenotype typically associated with biallelic *RECQL4* defects (PP4).

### Other findings in childhood onset genes with unrelated tumours and actionability

Germline alteration associated with childhood-onset CPS but identified in patients with tumours unrelated to the corresponding CPS included *CDKN2A*, *POLE,* and *FH* in Ewing sarcoma; *LZTR1*, *SRP72*, and *ELP1* in B-ALL; *KDM3B* in neuroblastoma; *NRAS* in paraganglioma with a concomitant *VHL* PGV; and *ELP1* in a rhabdoid tumour with a concomitant *SMARCB1* PGV. All findings were actionable except those involving *ELP1* and *KDM3B* (7/9 PGVs).

**Other findings - reduced-penetrance and risk alleles**

Reduced-penetrance (*CHEK2* p.Ile157Thr, n=2) and risk alleles (*APC* p.Ile1307Lys, n=3; *MITF* p.Glu318Lys, n=3) were classified as such in accordance with recently published consensus frameworks (8,9).

**References**


1. Kato S, Han SY, Liu W, Otsuka K, Shibata H, Kanamaru R, et al. Understanding the function-structure and function-mutation relationships of p53 tumor suppressor protein by high-resolution missense mutation analysis. Proc Natl Acad Sci U S A. 2003 Jul 8;100(14):8424–9.
2. Zerdoumi Y, Lanos R, Raad S, Flaman JM, Bougeard G, Frebourg T, et al. Germline TP53 mutations result into a constitutive defect of p53 DNA binding and transcriptional response to DNA damage. Hum Mol Genet. 2017 Jul 15;26(14):2812.
3. Wasserman JD, Novokmet A, Eichler-Jonsson C, Ribeiro RC, Rodriguez-Galindo C, Zambetti GP, et al. Prevalence and functional consequence of TP53 mutations in pediatric adrenocortical carcinoma: a children's oncology group study. J Clin Oncol. 2015 Feb 20;33(6):602–9.
4. Kotler E, Shani O, Goldfeld G, Lotan-Pompan M, Tarcic O, Gershoni A, et al. A Systematic p53 Mutation Library Links Differential Functional Impact to Cancer Mutation Pattern and Evolutionary Conservation. Mol Cell. 2018 Jul 5;71(1):178-190.e8.
5. Giacomelli AO, Yang X, Lintner RE, McFarland JM, Duby M, Kim J, et al. Mutational processes shape the landscape of TP53 mutations in human cancer. Nat Genet. 2018 Oct;50(10):1381–7.
6. Kawaguchi T, Kato S, Otsuka K, Watanabe G, Kumabe T, Tominaga T, et al. The relationship among p53 oligomer formation, structure and transcriptional activity using a comprehensive missense mutation library. Oncogene. 2005 Oct 20;24(46):6976–81.

7. Gentile M, Bergman Jungeström M, Olsen KE, Söderkvist P, Wingren S. p53 and survival in early onset breast cancer: analysis of gene mutations, loss of heterozygosity and protein accumulation. Eur J Cancer. 1999 Aug;35(8):1202–7.
8. Garrett A, Allen S, Durkie M, Burghel GJ, Robinson R, Callaway A, et al. Classification of variants of reduced penetrance in high-penetrance cancer susceptibility genes: Framework for genetics clinicians and clinical scientists by CanVIG-UK (Cancer Variant Interpretation Group-UK). Genet Med. 2025 Feb;27(2):101305.
9. Schmidt RJ, Steeves M, Bayrak-Toydemir P, Benson KA, Coe BP, Conlin LK, et al. Recommendations for risk allele evidence curation, classification, and reporting from the ClinGen Low Penetrance/Risk Allele Working Group. Genet Med. 2024 Mar;26(3):101036.
