## Supplementary tables legend for "Germline sequencing in children with cancer in Quebec: an integrated investigative approach"

**Table S1. Virtual 352 gene panel.**

**Table S2. Variant interpretation details.**

**Table S2A. ClinGen Expert Panel Specifications considered in the present study.**

Gene-specific ACMG/AMP specifications developed by ClinGen Expert Panels were systematically applied when available. The table lists all Expert Panel frameworks considered, with their latest version and release date at the time of variant interpretation.

**Table S2B. Adaptation of ACMG criteria for the present study in the absence of ClinGen Expert Panel Specifications.**

When no gene-specific ClinGen specification was available, ACMG/AMP 2015 criteria were applied with selected adaptations from recent expert recommendations. Adaptations mainly concerned PVS1, PS4, PM2, PM3, PP3, PP4, and PP5 as detailed in the table below.

References are indicated by superscript numbers within the table.

**Abbreviations:** AD, autosomal dominant; AR, autosomal recessive.

**Symbols:** "(" and ")" indicate exclusion of the end value and "[" and "]" indicate inclusion of the end value.

**References**


1. Abou Tayoun AN, Pesaran T, DiStefano MT, Oza A, Rehm HL, Biesecker LG, Harrison SM; ClinGen Sequence Variant Interpretation Working Group (ClinGen SVI). *Recommendations for interpreting the loss of function PVS1 ACMG/AMP variant criterion.* Hum Mutat. 2018;39(11):1517-1524.
2. Feurstein S, Hahn CN, Mehta N, Godley LA. *A practical guide to interpreting germline variants that drive hematopoietic malignancies, bone marrow failure, and chronic cytopenias.* Genet Med. 2022;24(4):931-954.
3. Pejaver V, Byrne AB, Feng BJ, Pagel KA, Mooney SD, Karchin R, O'Donnell-Luria A, Harrison SM, Tavtigian SV, Greenblatt MS, Biesecker LG, Radivojac P, Brenner SE. *Calibration of computational tools for missense variant pathogenicity classification and ClinGen recommendations for PP3/ BP4 criteria.* Am J Hum Genet. 2022;109(12):2163-2177.
4. Biesecker LG, Harrison SM; ClinGen Sequence Variant Interpretation Working Group. *The ACMG/AMP reputable source criteria for the interpretation of sequence variants.* Genet Med. 2018;20(12):1687-1688.

**Table S2C. Germline variants retained as pathogenic, likely pathogenic, reduced-penetrance or risk alleles in the present study.**

Variants were classified following ACMG/AMP guidelines, applying the corresponding ClinGen Expert Panel Specifications (Table S2A) or, when unavailable, the adapted ACMG framework detailed in Table S2B. *CHEK2* c.470T>C (p.Ile157Thr) was classified as *likely pathogenic – reduced penetrance* as per the CanVIG-UK framework for high-penetrance genes[1]. *APC* c.3920T>A (p.Ile1307Lys) and *MITF* c.1273G>A (p.Glu318Lys) were classified as *risk alleles* as per the ClinGen Low Penetrance/Risk Allele Working Group[2]. Three additional *VUS* were retained for downstream analyses based on strong supporting evidence, as described in the Methods.

**Abbreviations:** ACMG, American College of Medical Genetics and Genomics; DP, read depth; HGVSc, Human Genome Variation Society coding representation; HGVSg, Human Genome Variation Society genomic representation; HGVSp, Human Genome Variation Society protein representation; LP, likely pathogenic; P, pathogenic; VAF, variant allele frequency, VUS+, highly suspicious variant of uncertain significance.

**References:**


1. Garrett A, Allen S, Durkie M, Burghel GJ, Robinson R, Callaway A, *et al.* Classification of variants of reduced penetrance in high-penetrance cancer susceptibility genes: Framework for genetics clinicians and clinical scientists by CanVIG-UK (Cancer Variant Interpretation Group-UK). *Genet Med.* 2025;27(2):101305.
2. Schmidt RJ, Steeves M, Bayrak-Toydemir P, Benson KA, Coe BP, Conlin LK, *et al.* Recommendations for risk allele evidence curation, classification, and reporting from the ClinGen Low Penetrance/Risk Allele Working Group. *Genet Med.* 2024;26(3):101036.


**Table S3. Summary of the cohort.**

This table summarises the clinical and molecular characteristics of the 484 patients and their relevance to the cancer phenotype, including germline findings (GF), ACMG classification, zygosity, detection method, somatic second hits (for tumor suppressor gene), category of GF (diagnostic finding vs other finding) and the strength of association with the cancer phenotype if diagnostic finding (established vs suspicious), clinical actionability, and cancer predisposition syndrome (CPS) diagnosis.

**Abbreviations:** ACC, adrenocortical carcinoma; ALCL, anaplastic large cell lymphoma; aRMS, alveolar rhabdomyosarcoma; A-T, ataxia–telangiectasia; B-ALL, B-cell acute lymphoblastic leukemia; AMKL, acute megakaryoblastic leukemia; AML, acute myeloid leukaemia; APML, acute promyelocytic leukemia; B-NHL, B-cell non-Hodgkin lymphoma; Bone/STS, bone and soft tissue sarcoma; cBWS, clinical diagnosis of Beckwith–Wiedemann syndrome; CCHS, congenital central hypoventilation syndrome; CML, chronic myeloid leukemia; CPS, cancer predisposition syndrome; CRC, colorectal carcinoma; DF, diagnostic finding; DLBCL, diffuse large B-cell lymphoma; eRMS, embryonal rhabdomyosarcoma; EWS, Ewing sarcoma; FAMMM, familial atypical multiple mole melanoma syndrome; F, female; GCT, germ cell tumor; GF, germline finding; GI, gastrointestinal; HBOC, hereditary breast and ovarian cancer; Haem, haematologic neoplasm; HL, Hodgkin lymphoma; Hmz, homozygous; HLRCC, hereditary leiomyomatosis and renal cell cancer; HPT-JT, hyperparathyroidism–jaw tumour syndrome; Htz, heterozygous; JGCT, juvenile granulosa cell tumor; JMML, juvenile myelomonocytic leukemia; LBCL, large B-cell lymphoma; LFS, Li-Fraumeni syndrome; M, male; MDS, myelodysplastic syndrome; NA, not applicable; NBL, neuroblastoma; NET, neuroendocrine tumor; NF1, neurofibromatosis type 1; oSCST, ovarian sex cord–stromal tumor; oSLCT, ovarian Sertoli–Leydig cell tumor; OST, osteosarcoma; PEComa, perivascular epithelioid cell tumor; Ped, pediatric; P/LP, pathogenic or likely pathogenic variant; PNET, primitive neuroectodermal tumor; PPAP, polymerase proofreading-associated polyposis; PPB, pleuropulmonary blastoma; PPGL, pheochromocytoma/paraganglioma; RCC, renal cell carcinoma; RMS, rhabdomyosarcoma; RTPS-1, Rothmund–Thomson syndrome type 1; RTPS-2, Rothmund–Thomson syndrome type 2; T21, trisomy 21; T-ALL, T-cell acute lymphoblastic leukemia; TMB, tumour mutational burden; T-NHL, T-cell non-Hodgkin lymphoma; TSG, tumor suppressor gene; solid, solid cancer; VHL, von Hippel–Lindau disease; VUS+, highly suspicious variant of uncertain significance; WES, whole-exome-sequencing; XP, xeroderma pigmentosum.

**Table S4. Summary of second hits in patients with monoallelic diagnostic findings in tumour suppressor genes.**

**Notes:** 1: No reliable tumour WES data available for this patient.

**Abbreviations:** ACC, adrenocortical carcinoma; AR, autosomal recessive; DNE, dominant-negative effect; Htz, heterozygous; LOH, loss of heterozygosity; NA, not applicable; P/LP, pathogenic/likely pathogenic; PNET, primitive neuroectodermal tumour; RCC, renal cell carcinoma; RMS, rhabdomyosarcoma; VUS+, highly suspicious variant of uncertain significance.

**Table S5. Summary of splicing variants and their association to the cancer phenotype.**

**Abbreviations:** DF, diagnostic finding; HGVSc, Human Genome Variation Society coding representation

**Table S6. Summary of CPS and phenotype-genotype correlation.**
**Notes:** 1: only one of two P/LP variants identified in this patient with clear ataxia-telangiectasia phenotype

**Abbreviations:** APL, acute promyelocytic leukaemia; ASD, atrial septal defect; BWS, clinical diagnosis of Beckwith–Wiedemann syndrome; CPS, cancer predisposition syndrome; eRMS, embryonal rhabdomyosarcoma; F, female; hmz, homozygous; htz, heterozygous; JMML, juvenile myelomonocytic leukaemia; LOH, loss of heterozygosity; M, male; MMR, mismatch repair; NA, not applicable; NOS, not otherwise specified; PNET, primitive neuroectodermal tumor; P/LP, pathogenic or likely pathogenic variant; T21, trisomy 21; TMB, tumor mutational burden; VSD, ventricular septal defect; VUS+, highly suspicious variant of uncertain significance.
